# Deep Phenotyping Long-COVID Postural Tachycardia Syndrome

**DOI:** 10.1101/2025.04.28.25326587

**Authors:** Nicholas W. Larsen, Hash Brown Taha, Jordan Seliger, Ruba Shaik, Jannika Machnik, Maarten G. Lansberg, Srikanth Muppidi, Safwan S. Jaradeh, Christopher H. Gibbons, Paul J. Utz, Mitchell G. Miglis

**Author notes:** Corresponding Author: Mitchell G. Miglis, MD 213 Quarry Rd, Palo Alto, CA 94304, USA, M.

## Abstract

**Background & Objectives:** Long COVID postural tachycardia syndrome (LC-POTS) is increasingly recognized as a manifestation of post-viral autonomic dysfunction following SARS-CoV-2 infection, however its pathogenic mechanisms remain incompletely understood. We aimed to evaluate patterns of autonomic dysfunction in a cohort of carefully phenotyped, previously healthy participants with LC-POTS using autonomic, cerebrovascular, respiratory, blood, and tissue analyses. Comparison groups included participants with non-COVID POTS (NC-POTS) and controls with a history of SARS-CoV-2 infection without lasting sequaelae.

**Methods:** Participants completed a battery of autonomic function tests including measures of sudomotor, cardiovagal, and sympathetic adrenergic function, and head-up tilt (HUT) with transcranial Doppler measures of cerebral blood flow velocity (CBFv), end-tidal CO 2 (ETCO_2_), cerebral and skeletal muscle near-infrared spectroscopy (NIRS) and plasma catecholamines. Skin biopsy was performed at proximal and distal sites and analyzed for intraepidermal nerve fiber density (IENFD) and phosphorylated alpha-synuclein (P-Syn).

**Results:** The final cohort included 43 participants with LC-POTS, 17 with NC-POTS, and 27 controls. Both LC-POTS and NC-POTS had higher ΔHRs on HUT compared to controls (42 ± 18.6, 46 ± 15.9, 25 ± 12.9, p<0.01). 42.5% of LC-POTS had hyperadrenergic responses on HUT, however mean norepinephrine values were similar to NC-POTS and controls. LC-POTS participants had the lowest mean ETCO values of all groups at end-tilt (22.1 ± 7.5, 25.3 ± 5.8, 29.7 ± 7.3, p<0.01). While LC-POTS exhibited abnormalities across sudomotor (39%), sympathetic adrenergic (11.3%) and cardiovagal (9.3%) domains, we well as reduced IENFD (12.2%), there was no difference when compared to NC-POTS and controls. Measures of cerebral perfusion and oxygen extraction (vCBF, NIRS), were also similar. Dermal P-Syn deposition was identified in 5% of LC-POTS participants and absent in NC-POTS and controls.

**Conclusions:** Orthostatic tachycardia, sympathetic adrenergic hyperactivity, hypocapnia and other autonomic abnormalties were relatively common in our LC-POTS cohort, although many of these findings overlapped with NC-POTS and controls. Measures of cerebrovascular changes during orthostatic challenge were also similar and failed to correlate with symptoms of orthostatic intolerance.

## Introduction

While most individuals with coronavirus infectious disease 2019 (COVID-19) will recover without lasting sequelae, there is a growing population of those with chronic post-infectious symptoms, a term alternatively referred to as Long-COVID (LC) and post-acute sequelae of severe acute respiratory syndrome coronavirus-2 (PASC).^1^ Many symptoms of LC, including fatigue, lightheadedness, palpitations, chest pain, brain fog, and gastrointestinal symptoms, were observed in individuals with postural tachycardia syndrome (POTS) before the pandemic.^2^ POTS is defined by chronic orthostatic intolerance and an exaggerated postural tachycardia (sustained increase in heart rate (HR) by at least 30 beats per minute (bpm) in adults) in the absence of orthostatic hypotension.^3^ While POTS remains a syndrome with incompletely understood pathophysiological mechanisms, viral infection is the most commonly reported trigger of symptom onset, affecting by some estimates 50% of patients.^4^ Long-COVID POTS (LC-POTS) is being described with increasing frequency, and autonomic dysfunction is being increasingly recognized as a core feature of chronic post-viral syndromes including LC.^5–7^

Despite these associations, and the fact that POTS is a relatively common and disabling condition,^4^ the pathogenic mechanisms of POTS have remained incompletely understood for decades. The COVID-19 pandemic has thus created an opportunity to better understand the mechanisms of POTS and post-viral autonomic dysfunction with greater precision, given the known pathogen of severe acute respiratory syndrome-coronavirus-2 (SARS-CoV-2), large proportion of the general population who were infected, and availability of biospecimens. The goal of this study was to evaluate the type and severity of autonomic dysfunction in a subset of carefully phenotyped, previously healthy patients with LC-POTS using a detailed protocol of autonomic, cerebrovascular, respiratory, blood, and tissue analyses, utilizing non-COVID POTS (NC-POTS) and community controls as comparison groups.

## Methods

Adult participants with LC-POTS were consecutively recruited from the Stanford Autonomic Disorders Clinic between September 2021 and May 2025. Participants were diagnosed with LC-POTS by clinical autonomic neurologists with expertise in POTS (MM, NL, SM, SJ), based on symptoms of POTS that developed within one month of confirmed SARS-COV-2 infection and persisted for at least three months at the time of evaluation, with evidence of an exaggerated postural tachycardia on head-up tilt (HUT) testing or 10-minute active stand testing, according to current consensus criteria ^3^, prior to enrollment. All LC-POTS participants were healthy prior to SARS-CoV-2 infection without any significant past medical history that, in the investigator’s opinion, would predispose them to autonomic dysfunction. Controls were recruited by means of advertisements posted on the Stanford campus and social media outlets, and consisted of healthy participants with a history of confirmed SARS-CoV-2 infection ≥ 3 months from enrollment date without lasting sequelae. In addition, all controls were screened with a 10-minute active stand test to ensure absence of exaggerated postural tachycardia. A second comparison group of NC-POTS consisted of participants who developed POTS from non-COVID etiologies, all of whom were established and well-phenotyped patients of the investigators prior to study enrollment. SARS-CoV-2 infection was confirmed by real time-reverse transcriptase polymerase chain reaction (RT-PCR) or rapid antigen testing (RAT).

Exclusion criteria included age < 18 years (all groups), documented SARS-CoV-2 infection or symptoms of SARS-CoV-2 infection within 3 months of POTS symptom onset (NC-POTS), POTS symptom duration < 3 months (LC-POTS and NC-POTS), pregnancy or other comorbid conditions that could mimic POTS (LC-POTS and NC-POTS), and pre-existing conditions or symptoms suggestive of autonomic dysfunction prior to SARS-CoV-2 infection (LC-POTS).

All participants completed a battery of standardized autonomic reflex tests, including sudomotor measures (Quantitative Sweat Measurement System (Q-sweat), WR Medical Electronics Co., Stillwater, MN), parasympathetic cardiovagal measures (heart rate variability with deep breathing (HRDB), Valsalva HR ratio) and sympathetic adrenergic measures (Valsalva BP response, 10-minute HUT at an angle of 70 degrees). Beat-to-beat blood pressure (BP) was measured with finger plethysmography using a CNAP^®^ device (CN Systems Medizintechnik GmbH, Graz, Austria) and confirmed with an automated cuff sphygmomanometer over the brachial artery during HUT. Delta (Δ) HR was calculated as the difference between maximum and supine baseline HR measurements during HUT. Δ systolic BP (SBP) was calculated as the difference between supine baseline and minimum SBP measurements during HUT. The Composite Autonomic Severity Score (CASS), a quantitative measure of autonomic failure, was calculated for all autonomic reflex tests, as previously described.^8^

To further evaluate potential mechanisms of orthostatic intolerance in LC-POTS, additional neurophysiological modalities were utilized during HUT in all participants. To evaluate the role of cerebral hypoperfusion, cerebral blood flow velocity (CBFv) was measured with transcranial Doppler (TCD) probes positioned over bilateral middle cerebral arteries (MCAs) using a robotic headset (Nexgen Neurovision Robotic TCD System, Multigon Industries, Inc., Elmsford, NY; Dolphin/MAX, VIASONIX Vascular, Inc., Hixon, Tennessee). Δ CBFv values were calculated as the change in CBFv from the supine baseline to the final minute of HUT. Peak systolic velocity (PSV), end diastolic velocity (EDV), and the pulsatile index (PI) were also recorded. To evaluate the role of hypocapnia, simultaneous end-tidal carbon dioxide (ETCO_2_) measurements were obtained using nasal capnography with an oral scoop (Capnostream 35, Medtronics, Inc., Minneapolis, MN). Respiratory rate was recorded with a thoracic strain gauge and peripheral oxygen saturation (SpO_2_) with pulse oximeter. To evaluate the role of impaired cerebral and peripheral skeletal muscle oxygen extraction, simultaneous measurement of cerebral and skeletal muscle oxygen extraction was obtained with near-infrared spectroscopy (NIRS) sensors positioned over the right and left forehead and right gastrocnemius muscle (SenSmart Model X-100, Nonin Medical, Inc., Plymouth, MN). The Δ for all additional neurophysiological variables during HUT were calculated as the difference between supine baseline and the last minute of HUT. If participants exhibited a sudden hypotensive response during HUT consistent with vasovagal presyncope, subjects were returned to the supine position prior to syncope and two minutes prior to the hypotensive response was used as the participant’s last minute of HUT, to ensure that end-tilt CBFv was not influenced by tilt-evoked, neurally-mediated hypotension. Data was excluded for participants who failed to complete at least three minutes of HUT due to tilt-evoked, neuraly-mediated hypotensive responses.

To evaluate the role of excessive orthostatic norepinephrine (NE) release, supine and upright NE levels were obtained during HUT. A peripheral intravenous catheter (PIVC) was placed in the antecubital vein in the supine position. After PIVC placement, the participant remained in the supine position for ten minutes before a baseline NE sample was obtained, followed by a second NE sample obtained during the final minute of HUT. A hyperadrenergic response was defined as an upright NE level > 600 pg/ml or 3X greater than the baseline supine value, as previously described.^9^

To evaluate the role of peripheral autonomic denervation, participants underwent 3 mm punch biopsies at the distal leg (10 cm proximal from the medial malleolus), proximal thigh (10 cm proximal from the lateral knee), and cervical (3 cm lateral to C7 spinous process) regions. Biopsies were processed and analyzed by Cutaneous NeuroDiagnostics Life Sciences (Scottsdale, AZ) for intraepidermal nerve fiber density (IENFD) as well as phosphorylated alpha-synuclein, an exploratory measure, using immunohistochemistry techniques previously described.^10^ A multinational age- and gender-adjusted dataset was utilized for normative IENFD values.^11^ Additionally, a subset of participants completed olfactory testing with the University of Pittsburg Smell Idenfitication Test (UPSIT)-40 and underwent blood-based biomarker assays including hematologic, metabolic, inflammatory, coagulation, and immune profiling analyses. A set of serum and plasma samples were collected from all participants and biobanked for future analyses.

The combined cohort consisted of prospectively enrolled participants from two sequential studies. The first study was funded by Dysautonomia International and recruited 23 LC-POTS and 7 controls. The second study was funded by the National Institutes under the RECOVER Pathobiology Program and recruited an additional 20 LC-POTS, 17 NC-POTS, and 20 controls. The studies were identical in methodology, with the exception of UPSIT-40 and serology measures, which were obtained in the RECOVER cohort only (Figure 1).

**Figure 1.**
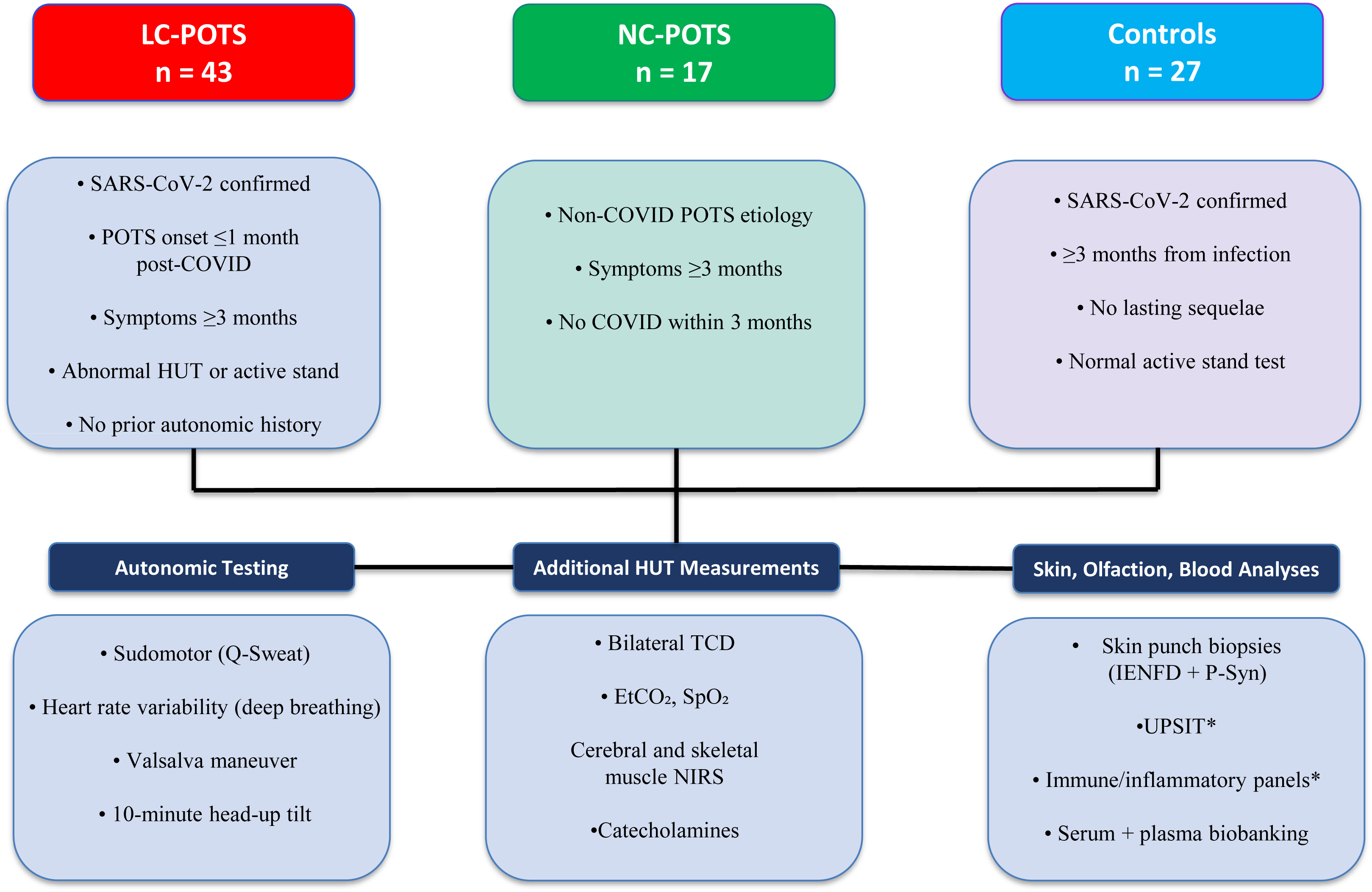
Study Design and Participant Flow. SARS-CoV-2 = Severe Acute Respiratory Syndrome-Coronavirus-2, POTS = postural tachycardia syndrome, COVID = coronavirus disease, HUT = head-uptilt, TCD = transcranial doppler, EtCO_2_ = end-tidal CO_2_, SpO^2^ = peripheral oxygen saturation, NIRS = near-infared spectroscopy, IENFD = intraepidermal nerve fiber density, UPSIT = University of Pennsylvania smell identification test.

### Statistical Analysis

Normality was assessed using the Shapiro–Wilk test. For three-group comparisons, normally distributed variables were analyzed using one-way analysis of variance (ANOVA) with Tukey’s honestly significant difference (HSD) test for post hoc pairwise comparisons, while non-normally distributed variables were analyzed using the Kruskal–Wallis test with pairwise Wilcoxon rank-sum tests and Benjamini–Hochberg adjustment. For pre- and post-omicron comparisons, normally distributed variables were analyzed using independent t-tests and non-normally distributed variables using Mann–Whitney U tests. Categorical variables were compared using Chi-square tests or Fisher’s exact tests when expected counts were <5. Pairwise post hoc comparisons for categorical variables were performed using pairwise proportion tests with Benjamini-Hochberg adjustment. Statistical significance was defined as a two-sided p < 0.05.**

The study was approved by the Stanford University Institutional Review Board (IRB-60096, IRB-69674) and all participants gave informed consent prior to participating.

## Results

The final cohort included 43 participants with LC-POTS, 17 particpants with NC-POTS, and 27 controls (Table 1). 6/17 (35%) of NC-POTS participants reported a trigger of symptom onset, including non-COVID viral illness (n=3), physical overexertion (n=2), and concussion (n=1). The remainder (65%) reported no discrete trigger of illness. LC-POTS participants were slightly older and had higher body mass index (BMI) compared to NC-POTS. The cohort was predominantly female in all groups with no significant differences. Racial distribution differed, with a higher percentage of White participants in the LC-POTS group compared to controls, as expected based on demographics reported in other POTS cohorts.^4^ The time from symptom onset (LC-POTS and NC-POTS) or infection (controls) to study date of autonomic testing was shorter in LC-POTS (20 ± 13.5 months vs 31.8 ± 18.2 in NC-POTS and 30.5 ± 13.9 in controls). LC-POTS participants were more likely to have hospitalized during the acute phase of their COVID-19 illness (16.3% vs 0% controls) and were more likely to be unvaccinated at the time of infection (53.5% vs 74.1% of controls). High rates of mild hyposmia were noted across groups (75% LC-POTS, 62.5% NC-POTS, 47.4% controls) without significant differences. Using the index date of December 1^st^, 2021 for the Omicron variant, as previously reported,^1^ 20 (46.6%) of LC-POTS participants were defined as pre-Omicron infections, and 23 (53.4%) were defined as post-Omicron infections, with no significant differences in demographics (Supplementary Table 1) except for vaccination at time of infection (20% fully vaccinated pre-omicron vs. 82.6% fully vaccinated post-omicron).

**Table 1.** Participant Demographics and Clinical Characteristics.

| Demographics | LC-POTS (n = 43) | NC-POTS (n = 17) | Control (n = 27) | P-value | Post-hoc |
| --- | --- | --- | --- | --- | --- |
| Age, mean (SD) | 34.1 (8.9) | 27.9 (8) | 30.5 (6.1) | 0.030 | LC- vs. NC-POTS |
| Gender, n (%) |  |  |  |  |  |
| Male | 5 (12) | 1 (6) | 9 (33) | 0.035 | NS |
| Female | 38 (88) | 16 (94) | 18 (67) |  |  |
| Race, n (%) |  |  |  |  |  |
| White | 34 (79) | 12 (71) | 13 (48) | 0.013 | LC-POTS vs. Control |
| White/Hispanic | 0 (0) | 0 (0) | 1 (4) |  |  |
| Black | 1 (2) | 0 (0) | 2 (7) |  |  |
| Asian | 5 (12) | 2 (12) | 8 (30) |  |  |
| Native Hawaiian/Other Pacific Islander | 1 (2) | 0 (0) | 0 (0) |  |  |
| Other | 0 (0) | 0 (0) | 3 (11) |  |  |
| Not disclosed | 2 (5) | 3 (18) | 0 (0) |  |  |
| BMI (kg/m<sup>2</sup>), mean (SD) | 27.4 (6.8) | 22.2 (3.9) | 24.8 (4.4) | 0.0082 | LC vs. NC-POTS |
| Time from infection to ANS testing (months), mean (SD) | 20 (13.5) | NA | 30.5 (13.9) | 0.0032 | — |
| COVID Severity, n (%) |  |  |  |  |  |
| Non-hospitalized | 36 (83.7) | NA | 27 (100) | 0.038 | — |
| Hospitalized | 7 (16.3) | NA | 0 (0) |  |  |
| Vaccinated at time of COVID infection, n (%) |  |  |  |  |  |
| Yes | 23 (53.5) | NA | 20 (74.1) | 0.14 | — |
| UPSIT-40<sup>I</sup> |  |  |  |  |  |
| Mean (SD) | 33.9 (2.6) | 34.7 (2.7) | 35.2 (2.9) | 0.33 | — |
| Abnormal, n (%) | 20 (75) | 16 (62.5) | 9 (47.4) | 0.22 | — |

Values for continuous variables are presented as mean (SD). Comparisons for continuous variables were performed using Kruskal-Wallis followed by posthoc pairwise Wilcoxon rank-sum tests with Benjamini-Hochberg adjustment or Mann-Whitney U tests as appropriate. Comparisons for categorical variables were performed using Chi-square tests for $\geq 5$ or Fisher's exact tests for $n < 5$ .
<sup>1</sup>data available for 20-LC POTS, 16 NC-POTS and 19 controls.
BMI = body mass index. ANS = autonomic nervous system; SD = standard deviation; COVID = coronavirus disease 2019; LC-POTS = Long-COVID postural tachycardia syndrome; NC-POTS = Non-COVID postural tachycardia syndrome; NS = non-significant.

Autonomic function testing results are summarized in Table 2. Valsalva ratio differed significantly across groups, with controls exhibiting lower mean values than both LC-POTS and NC-POTS, however rates of abnormal results were similar across groups. Sudomotor testing revealed significantly reduced Q-Sweat responses in both POTS groups at the forearm and proximal thigh, however no differences in the rates of abnormal results at any site across groups. No other significant differences were observed across other domains of cadiovagal or sympathetic adrenergic function, and CASS scores were similar across groups. While nearly half of LC- and NC-POTS participants had elevated NE levels on HUT, mean results were not statistically different from controls. The frequency and severity of autonomic abnormalities were similar across pre- and post-omicron LC-POTS symptom onset (Supplementary Table 2).

**Table 2.**
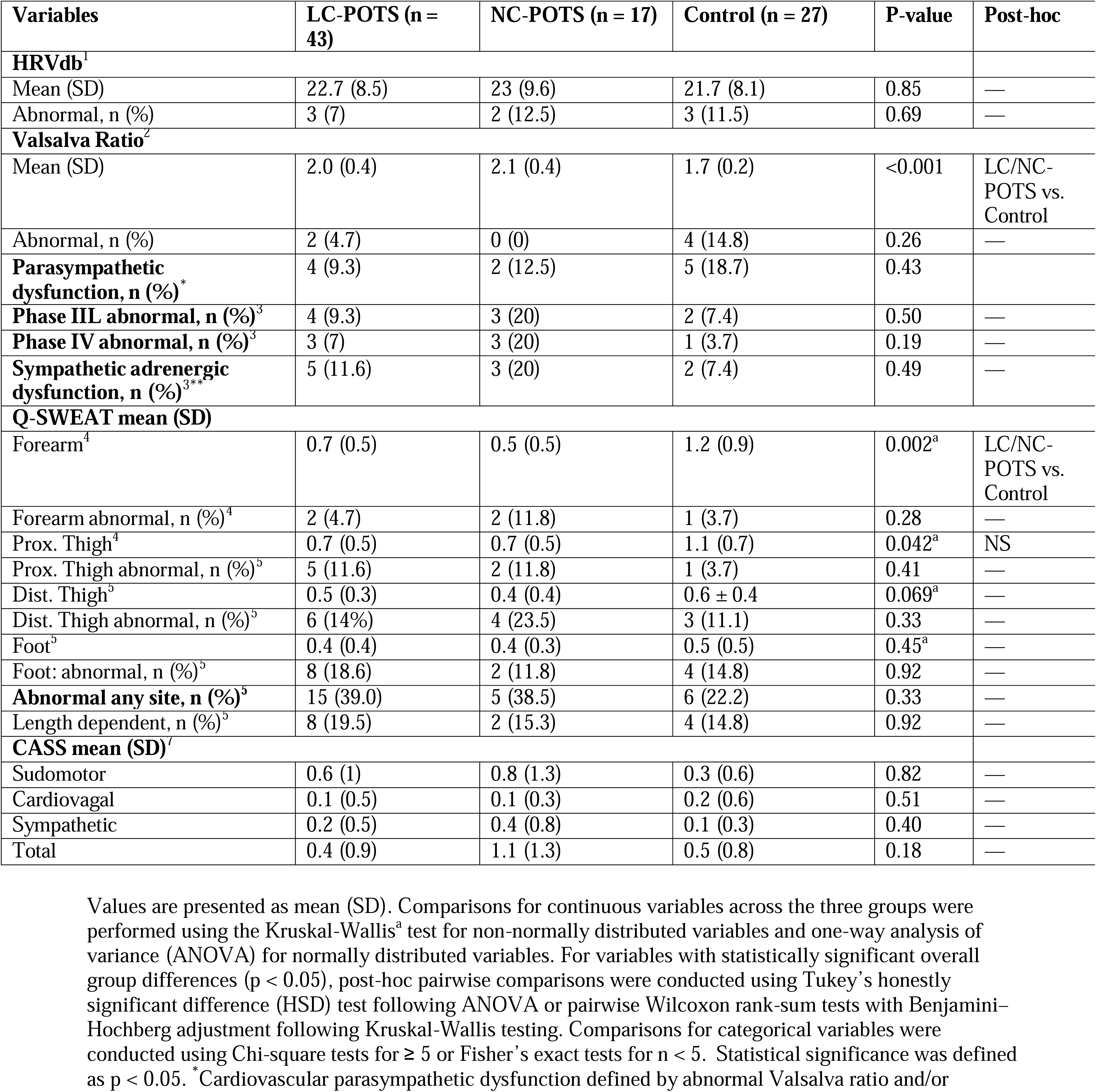
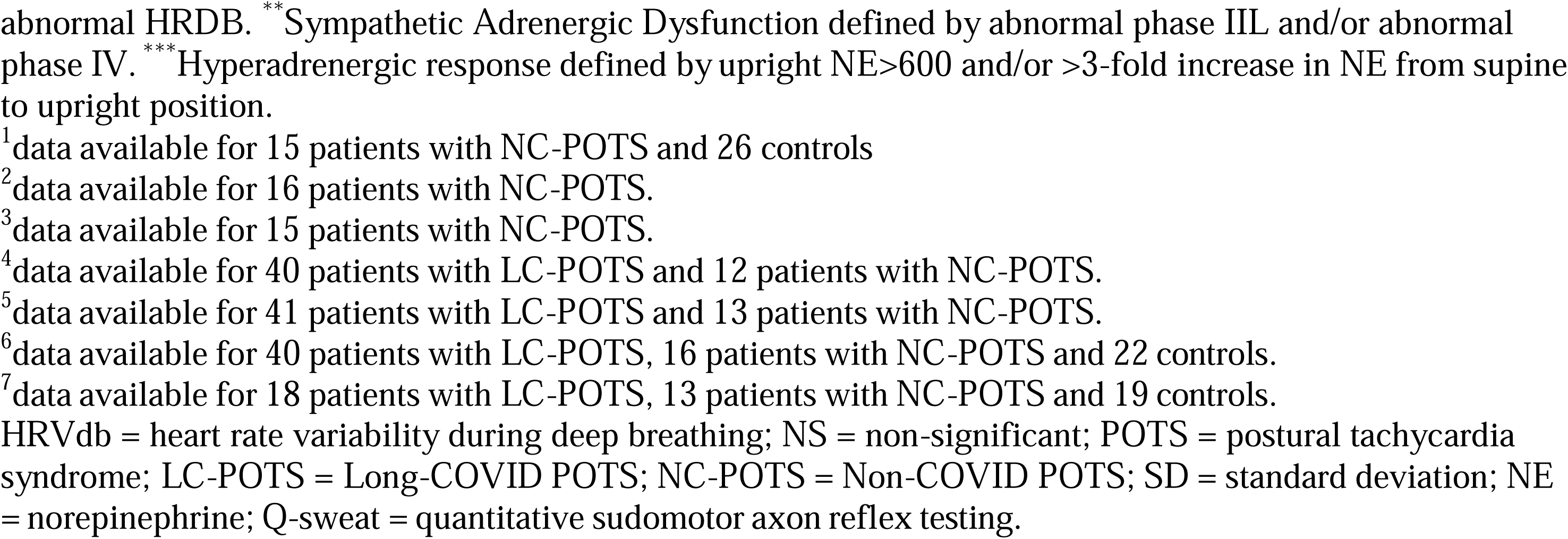
Autonomic Reflex Testing Results.

HUT data are summarized in Table 3. HR was higher in both LC-POTS and NC-POTS when compared to controls, as expected based on inclusion criteria, and LC-POTS participants had higher SBPs on HUT. LC-POTS participants had the lowest mean ETCO values of all groups at both supine baseline and end-tilt, with supine ETCO values significantly lower than NC-POTS, and end-tilt ETCO significantly lower than controls. The magnitude of ΔETCO and ΔRR (Supine - End-tilt) were greater in LC- and NC-POTS compared to controls. Peripheral oxygen saturation was normal in all groups. NIRS recording demonstrated lower supine skeletal muscle rSO in LC-POTS compared to controls. There was no difference in upright skeletal muscle or cerebral rSO among groups during HUT.

**Table 3.**
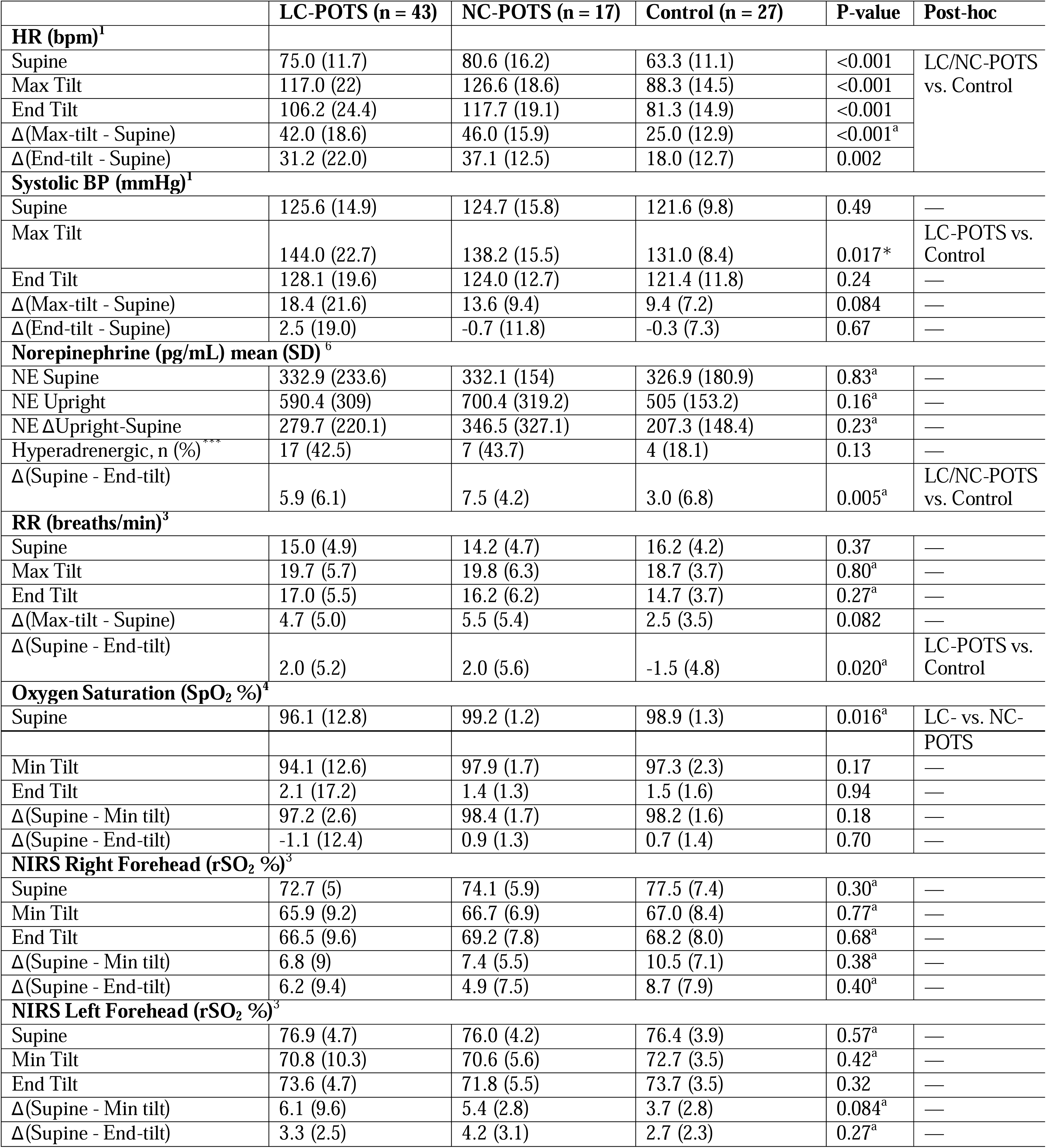
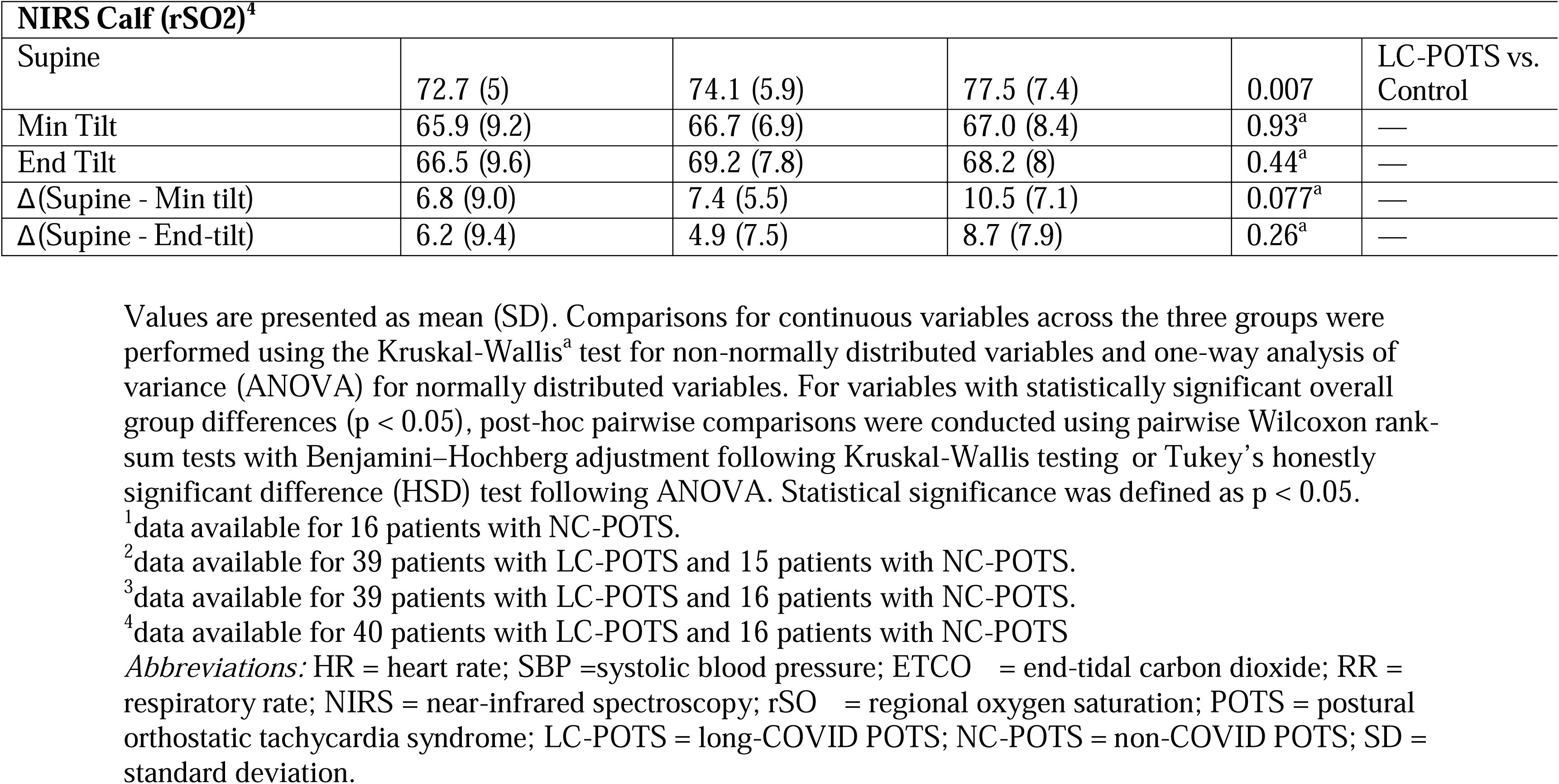
Head-Up Tilt Results.

|  | LC-POTS (n = 43) | NC-POTS (n = 17) | Control (n = 27) | P-value | Post-hoc |
| --- | --- | --- | --- | --- | --- |
| **HR (bpm)<sup>1</sup>** |  |  |  |  |  |
| Supine | 75.0 (11.7) | 80.6 (16.2) | 63.3 (11.1) | <0.001 | LC/NC-POTS<br>vs. Control |
| Max Tilt | 117.0 (22) | 126.6 (18.6) | 88.3 (14.5) | <0.001 |  |
| End Tilt | 106.2 (24.4) | 117.7 (19.1) | 81.3 (14.9) | <0.001 |  |
| Δ(Max-tilt - Supine) | 42.0 (18.6) | 46.0 (15.9) | 25.0 (12.9) | <0.001<sup>a</sup> |  |
| Δ(End-tilt - Supine) | 31.2 (22.0) | 37.1 (12.5) | 18.0 (12.7) | 0.002 |  |
| **Systolic BP (mmHg)<sup>1</sup>** |  |  |  |  |  |
| Supine | 125.6 (14.9) | 124.7 (15.8) | 121.6 (9.8) | 0.49 | — |
| Max Tilt | 144.0 (22.7) | 138.2 (15.5) | 131.0 (8.4) | 0.017* | LC-POTS vs.<br>Control |
| End Tilt | 128.1 (19.6) | 124.0 (12.7) | 121.4 (11.8) | 0.24 | — |
| Δ(Max-tilt - Supine) | 18.4 (21.6) | 13.6 (9.4) | 9.4 (7.2) | 0.084 | — |
| Δ(End-tilt - Supine) | 2.5 (19.0) | -0.7 (11.8) | -0.3 (7.3) | 0.67 | — |
| **Norepinephrine (pg/mL) mean (SD)<sup>6</sup>** |  |  |  |  |  |
| NE Supine | 332.9 (233.6) | 332.1 (154) | 326.9 (180.9) | 0.83<sup>a</sup> | — |
| NE Upright | 590.4 (309) | 700.4 (319.2) | 505 (153.2) | 0.16<sup>a</sup> | — |
| NE ΔUpright-Supine | 279.7 (220.1) | 346.5 (327.1) | 207.3 (148.4) | 0.23<sup>a</sup> | — |
| Hyperadrenergic, n (%)<sup>***</sup> | 17 (42.5) | 7 (43.7) | 4 (18.1) | 0.13 | — |
| Δ(Supine - End-tilt) | 5.9 (6.1) | 7.5 (4.2) | 3.0 (6.8) | 0.005<sup>a</sup> | LC/NC-POTS<br>vs. Control |
| **RR (breaths/min)<sup>3</sup>** |  |  |  |  |  |
| Supine | 15.0 (4.9) | 14.2 (4.7) | 16.2 (4.2) | 0.37 | — |
| Max Tilt | 19.7 (5.7) | 19.8 (6.3) | 18.7 (3.7) | 0.80<sup>a</sup> | — |
| End Tilt | 17.0 (5.5) | 16.2 (6.2) | 14.7 (3.7) | 0.27<sup>a</sup> | — |
| Δ(Max-tilt - Supine) | 4.7 (5.0) | 5.5 (5.4) | 2.5 (3.5) | 0.082 | — |
| Δ(Supine - End-tilt) | 2.0 (5.2) | 2.0 (5.6) | -1.5 (4.8) | 0.020<sup>a</sup> | LC-POTS vs.<br>Control |
| **Oxygen Saturation (SpO<sub>2</sub> %)<sup>4</sup>** |  |  |  |  |  |
| Supine | 96.1 (12.8) | 99.2 (1.2) | 98.9 (1.3) | 0.016<sup>a</sup> | LC- vs. NC-<br>POTS |
| Min Tilt | 94.1 (12.6) | 97.9 (1.7) | 97.3 (2.3) | 0.17 | — |
| End Tilt | 2.1 (17.2) | 1.4 (1.3) | 1.5 (1.6) | 0.94 | — |
| Δ(Supine - Min tilt) | 97.2 (2.6) | 98.4 (1.7) | 98.2 (1.6) | 0.18 | — |
| Δ(Supine - End-tilt) | -1.1 (12.4) | 0.9 (1.3) | 0.7 (1.4) | 0.70 | — |
| **NIRS Right Forehead (rSO<sub>2</sub> %)<sup>3</sup>** |  |  |  |  |  |
| Supine | 72.7 (5) | 74.1 (5.9) | 77.5 (7.4) | 0.30<sup>a</sup> | — |
| Min Tilt | 65.9 (9.2) | 66.7 (6.9) | 67.0 (8.4) | 0.77<sup>a</sup> | — |
| End Tilt | 66.5 (9.6) | 69.2 (7.8) | 68.2 (8.0) | 0.68<sup>a</sup> | — |
| Δ(Supine - Min tilt) | 6.8 (9) | 7.4 (5.5) | 10.5 (7.1) | 0.38<sup>a</sup> | — |
| Δ(Supine - End-tilt) | 6.2 (9.4) | 4.9 (7.5) | 8.7 (7.9) | 0.40<sup>a</sup> | — |
| **NIRS Left Forehead (rSO<sub>2</sub> %)<sup>3</sup>** |  |  |  |  |  |
| Supine | 76.9 (4.7) | 76.0 (4.2) | 76.4 (3.9) | 0.57<sup>a</sup> | — |
| Min Tilt | 70.8 (10.3) | 70.6 (5.6) | 72.7 (3.5) | 0.42<sup>a</sup> | — |
| End Tilt | 73.6 (4.7) | 71.8 (5.5) | 73.7 (3.5) | 0.32 | — |
| Δ(Supine - Min tilt) | 6.1 (9.6) | 5.4 (2.8) | 3.7 (2.8) | 0.084<sup>a</sup> | — |
| Δ(Supine - End-tilt) | 3.3 (2.5) | 4.2 (3.1) | 2.7 (2.3) | 0.27<sup>a</sup> | — |

| **NIRS Calf (rSO<sub>2</sub>)<sup>4</sup>** |  |  |  |  |  |
| --- | --- | --- | --- | --- | --- |
| Supine | 72.7 (5) | 74.1 (5.9) | 77.5 (7.4) | 0.007 | LC-POTS vs. Control |
| Min Tilt | 65.9 (9.2) | 66.7 (6.9) | 67.0 (8.4) | 0.93<sup>a</sup> | — |
| End Tilt | 66.5 (9.6) | 69.2 (7.8) | 68.2 (8) | 0.44<sup>a</sup> | — |
| Δ(Supine - Min tilt) | 6.8 (9.0) | 7.4 (5.5) | 10.5 (7.1) | 0.077<sup>a</sup> | — |
| Δ(Supine - End-tilt) | 6.2 (9.4) | 4.9 (7.5) | 8.7 (7.9) | 0.26<sup>a</sup> | — |

Values are presented as mean (SD). Comparisons for continuous variables across the three groups were performed using the Kruskal-Wallis<sup>a</sup> test for non-normally distributed variables and one-way analysis of variance (ANOVA) for normally distributed variables. For variables with statistically significant overall group differences ( $p < 0.05$ ), post-hoc pairwise comparisons were conducted using pairwise Wilcoxon rank-sum tests with Benjamini–Hochberg adjustment following Kruskal-Wallis testing or Tukey’s honestly significant difference (HSD) test following ANOVA. Statistical significance was defined as $p < 0.05$ .
<sup>1</sup>data available for 16 patients with NC-POTS.
<sup>2</sup>data available for 39 patients with LC-POTS and 15 patients with NC-POTS.
<sup>3</sup>data available for 39 patients with LC-POTS and 16 patients with NC-POTS.
<sup>4</sup>data available for 40 patients with LC-POTS and 16 patients with NC-POTS
*Abbreviations:* HR = heart rate; SBP =systolic blood pressure; ETCO<sub>2</sub> = end-tidal carbon dioxide; RR = respiratory rate; NIRS = near-infrared spectroscopy; rSO<sub>2</sub> = regional oxygen saturation; POTS = postural orthostatic tachycardia syndrome; LC-POTS = long-COVID POTS; NC-POTS = non-COVID POTS; SD = standard deviation.

TCD findings are summarized in Table 4. Values in LC-POTS, NC-POTS and controls, including right and left MCA CBFv, PSV, EDV, and PI during HUT were broadly similar, with no meaningful group-level differences observed, despite the fact that all POTS partipants noted severe symptoms of orthostatic intolerance during HUT, compared to mostly asymptomatic controls. Supine baseline to minute-by-minute HUT exploration of SBP, DBP, HR, ETCO , RR, SpO , rSO (right cerebral, left cerebral, and calf), and MCA CBFv (right, left and combined) showed clear group separation primarily in HR and ETCO (Figure 2) as LC-POTS and NC-POTS exhibited higher HR and lower ETCO compared to controls throughout HUT. Peak SBP demonstrated modest separation in LC-POTS compared to controls, while DBP and RR showed less consistent differences. In contrast, rSO and cerebral blood flow indices showed substantial overlap across groups over time, indicating no meaningful differences in cerebral perfusion and oxygen extraction dynamics.

**Figure 2.**
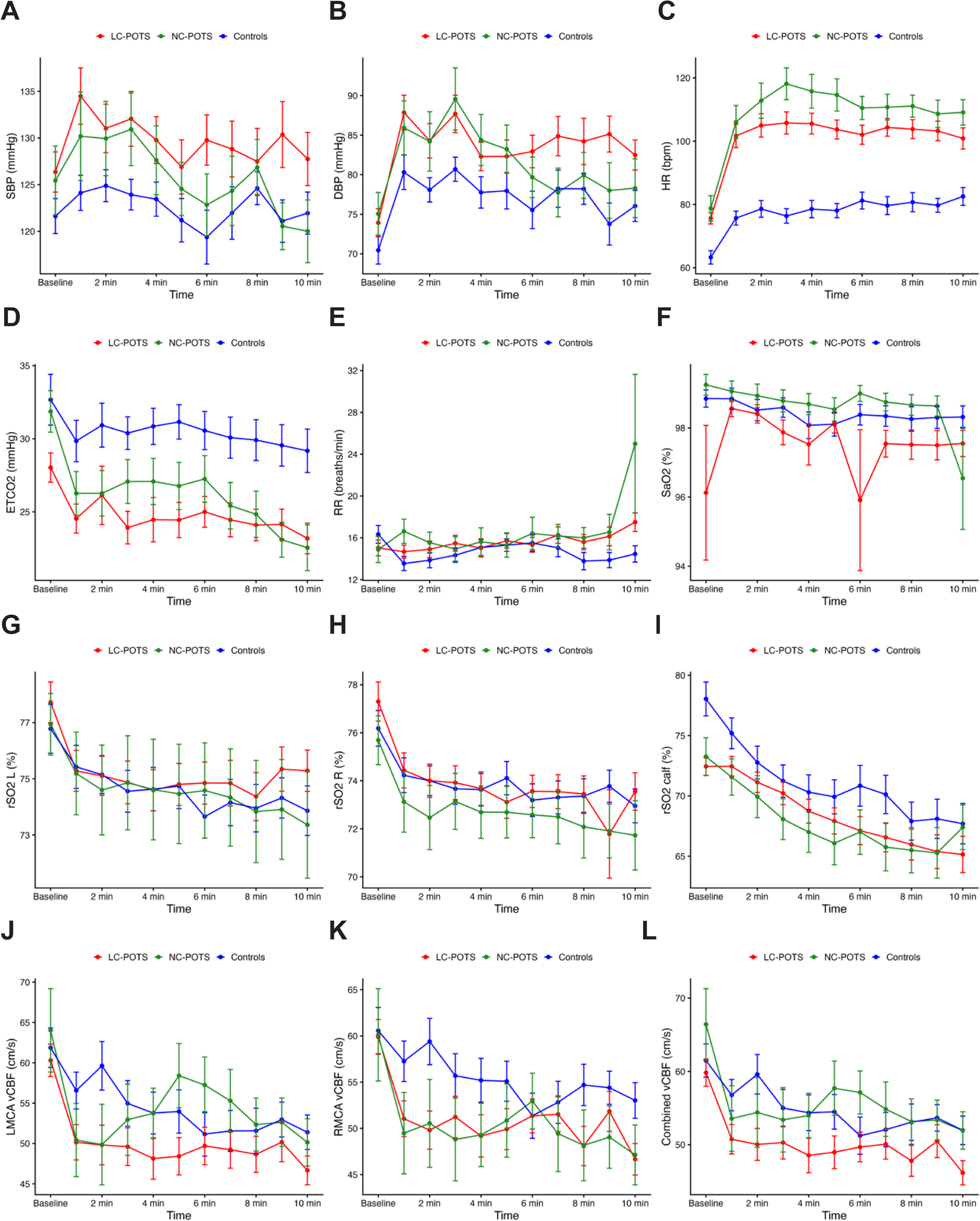
Physiological Responses and Cerebral Blood Flow Dynamics During Head-up Tilt Across Study Groups. Panels display mean ± SEM over time. (A) Systolic blood pressure, (B) Diastolic blood pressure, (C) Heart rate, (D) End-tidal carbon dioxide, (E) Respiratory rate, (F) Oxygen saturation (SaO ), (G) Left cerebral oxygen saturation (rSO -L), (H) Right cerebral oxygen extraction (rSO -R), (I) Calf oxygen extraction (rSO -calf), (J) Left middle cerebral artery cerebral blood flow velocity (LMCA CBFv), (K) Right middle cerebral artery cerebral blood flow velocity (RMCA CBFv), and (L) Combined middle cerebral artery cerebral blood flow velocity (MCA CBFv). *RECOVER cohort only.

**Table 4.** Head-Up Tilt Transcranial Doppler Results.

| **TCD Variable** | **LC-POTS (n = 32))** | **NC-POTS (n = 13)** | **Control (n = 26)** | **P-value** |
| --- | --- | --- | --- | --- |
| **Baseline Left MCA** |  |  |  |  |
| Mean vCBF (cm/s)<sup>1</sup> | 61.3 (12.6) | 64 (21.3) | 59.8 (14.7) | 0.74 |
| PSV (cm/s)<sup>1</sup> | 91.9 (19.0) | 91.5 (26.5) | 90.7 (23.1) | 0.98 |
| EDV (cm/s)<sup>2</sup> | 42.3 (7.9) | 44.5 (15.7) | 42.9 (9.1) | 0.85 |
| PI<sup>1</sup> | 0.8 (0.1) | 0.8 (0.2) | 0.8 (0.1) | 0.52 |
| **End Tilt Left MCA** |  |  |  |  |
| Mean vCBF (cm/s)<sup>3</sup> | 47 (14.7) | 46.8 (14.3) | 48.8 (12.9) | 0.87 |
| PSV (cm/s)<sup>3</sup> | 75 (23.1) | 72.4 (20.6) | 75.5 (18.6) | 0.91 |
| EDV (cm/s)<sup>2</sup> | 33.2 (9.3) | 33.3 (12) | 35.8 (10.5) | 0.70 |
| PI<sup>3</sup> | 0.9 (0.3) | 0.9 (0.2) | 0.8 (0.2) | 0.41<sup>a</sup> |
| **Δ Left MCA** |  |  |  |  |
| Mean vCBF (cm/s) | 14.4 (12.3) | 17.2 (11.4) | 10.9 (8.3) | 0.58 |
| PSV (cm/s) | 17 (13.6) | 19.1 (9.6) | 15.2 (11.3) | 0.34 |
| EDV (cm/s) | 9.1 (11.7) | 11.3 (9.7) | 7.1 (7.3) | 0.68 |
| PI | -0.1 (0.3) | -0.1 (0.2) | 0 (0.2) | 0.26<sup>a</sup> |
| **Baseline Right MCA** |  |  |  |  |
| Mean vCBF (cm/s)<sup>4</sup> | 60.3 (12.5) | 63.1 (19.8) | 59.8 (13.3) | 0.78 |
| PSV (cm/s)<sup>4</sup> | 91.6 (18.4) | 93.9 (22.6) | 90 (17.5) | 0.83 |
| EDV (cm/s)<sup>5</sup> | 43.1 (8.4) | 43.5 (15.5) | 42 (9.8) | 0.92 |
| PI<sup>4</sup> | 0.8 (0.2) | 0.8 (0.2) | 0.9 (0.2) | 0.89<sup>a</sup> |
| **End Tilt Right MCA** |  |  |  |  |
| Mean vCBF (cm/s)<sup>6</sup> | 50.2 (12.3) | 47.9 (11.8) | 51.5 (11.3) | 0.68 |
| PSV (cm/s)<sup>6</sup> | 80.9 (20.2) | 74.1 (16.6) | 80.3 (15.8) | 0.49 |
| EDV (cm/s)<sup>5</sup> | 35.6 (8.7) | 35 (9.2) | 38.1 (9.4) | 0.57 |
| PI<sup>6</sup> | 0.9 (0.3) | 0.8 (0.2) | 0.9 (0.2) | 0.57<sup>a</sup> |
| **Δ Right MCA** |  |  |  |  |
| Mean vCBF (cm/s) | 10.7 (9.8) | 15.2 (12.5) | 7.9 (8.7) | 0.11<sup>a</sup> |
| PSV (cm/s) | 11.7 (12.4) | 19.9 (12.2) | 10.4 (10.9) | 0.060 |
| EDV (cm/s) | 7.5 (8.6) | 8.5 (10.7) | 3.1 (8.2) | 0.18 |
| PI | -0.1 (0.3) | 0 (0.2) | 0 (0.2) | 0.19<sup>a</sup> |

Values are presented as mean (SD). Comparisons for continuous variables across the three groups were performed using the Kruskal-Wallis<sup>a</sup> test for non-normally distributed variables and one-way analysis of variance (ANOVA) for normally distributed variables. For variables with statistically significant overall group differences ( $p < 0.05$ ), post-hoc pairwise comparisons were conducted using Tukey's honestly significant difference (HSD) test following ANOVA or pairwise Wilcoxon rank-sum tests with Benjamini-Hochberg adjustment following Kruskal-Wallis testing. Statistical significance was defined as $p < 0.05$ .
<sup>1</sup>data available for 12 patients with NC-POTS and 22 controls.
<sup>2</sup>data available for 19 patients with LC-POTS, 12 patients with NC-POTS and 18 controls.
<sup>3</sup>data available for 31 patients with LC-POTS, 12 patients with NC-POTS and 22 controls.
<sup>4</sup>data available for 30 patients with LC-POTS and 25 controls.
<sup>5</sup>data available for 19 patients with LC-POTS and 21 controls.
<sup>6</sup>data available for 28 patients with LC-POTS.
TCD = transcranial Doppler; vCBF = cerebral blood flow velocity MCA = middle cerebral artery; PSV = peak systolic velocity; EDV = end diastolic velocity; PI = pulsatility index; POTS = postural tachycardia syndrome; LC-POTS = Long-COVID POTS; NC-POTS = Non-COVID POTS; SD = standard deviation.

Skin biopsy findings are summarized in Table 5. IENFD differed across groups, with higher values in NC-POTS compared to both LC-POTS and controls at the distal thigh and distal leg, and at the cervical site compared to controls, while no meaningful differences were observed between LC-POTS and controls. The percentage of participants with reduced IENFD at any site were low and similar across groups. Dermal P-Syn deposition was seen in 2 LC-POTS participants (5%, Figure 3) and was absent in NC-POTS and controls, with no significant differences between groups. Frequency and severity of skin biopsy abnormalities were similar across pre- and post-omicron LC-POTS symptom onset (Supplementary Table 5).

**Figure 3.**
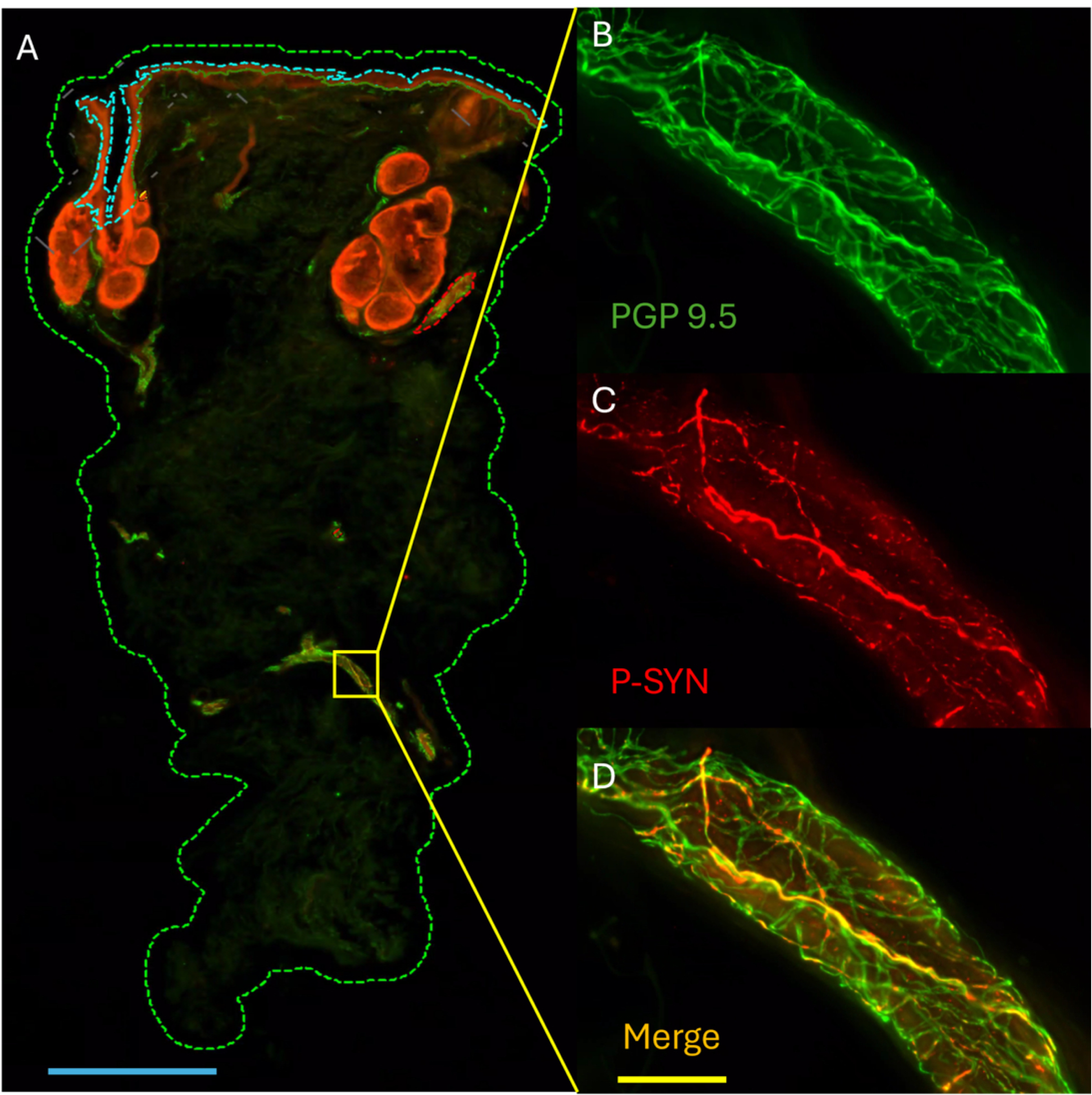
Cutaneous Immunostained Region of Skin. In A, a skin biopsy (outlined in dotted green to aid in visualization) is shown with the epidermis at the top (with a dotted blueline for visualization), and the deeper dermal structures seen beneath including hair follicles, sebaceous glands, pilomotor muscles and blood vessels. A blood vessel is outlined in yellow and can be viewed in images B-D. In B, a blood vessel immunostained with protein gene product 9.5 (PGP9.5) in green. In C, the same blood vessel is immunostained with phosphorylated alpha-synuclein (P-SYN) in red. In D, the merged image is seen confirming the presence of intra-axonal phosphorylated alpha-synuclein (seen in yellow/orange). Blue scale bar (in image A) = 1 millimeter. Yellow scale bar = 50 micrometers for images B-D.

**Table 5.**
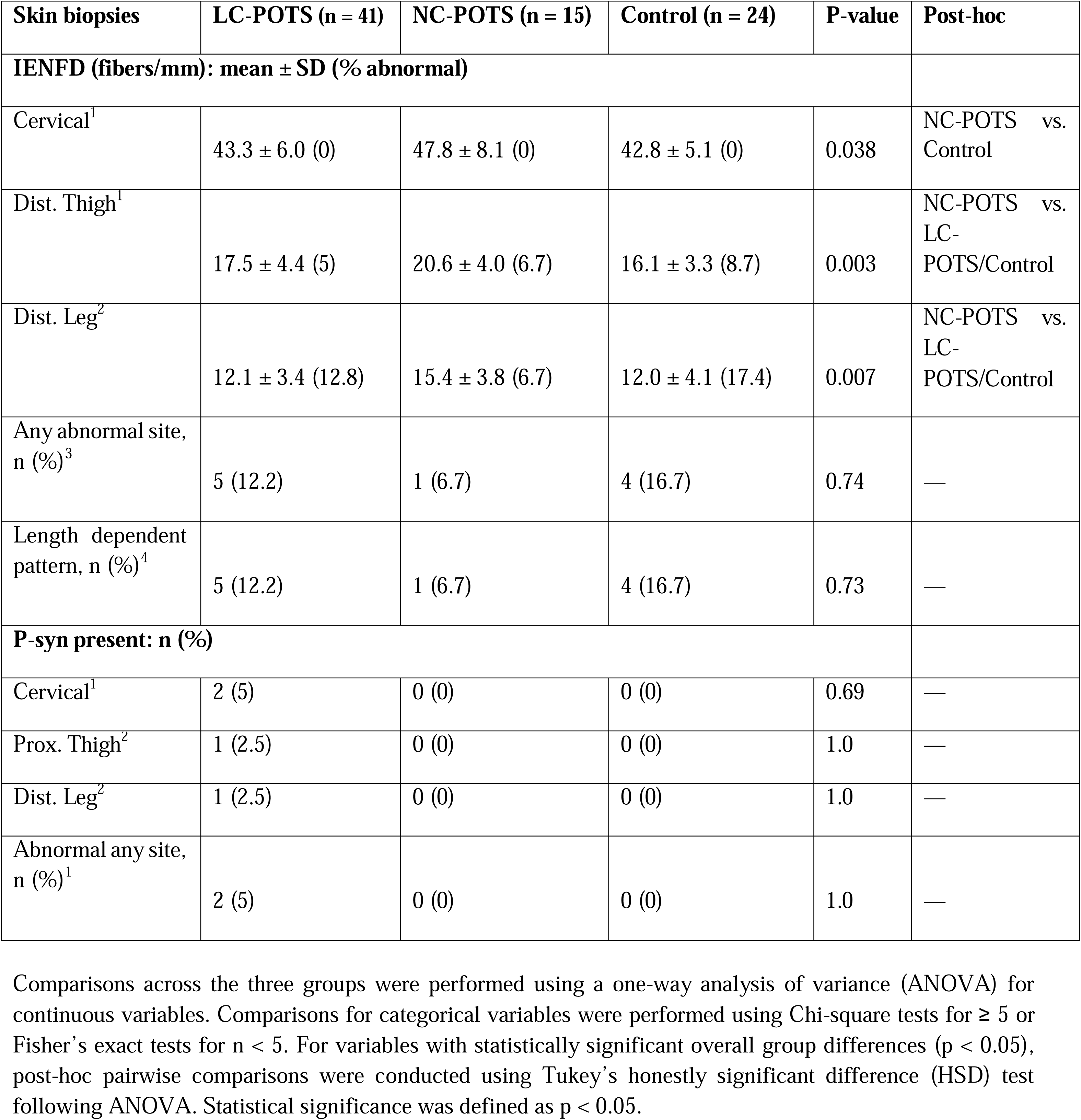
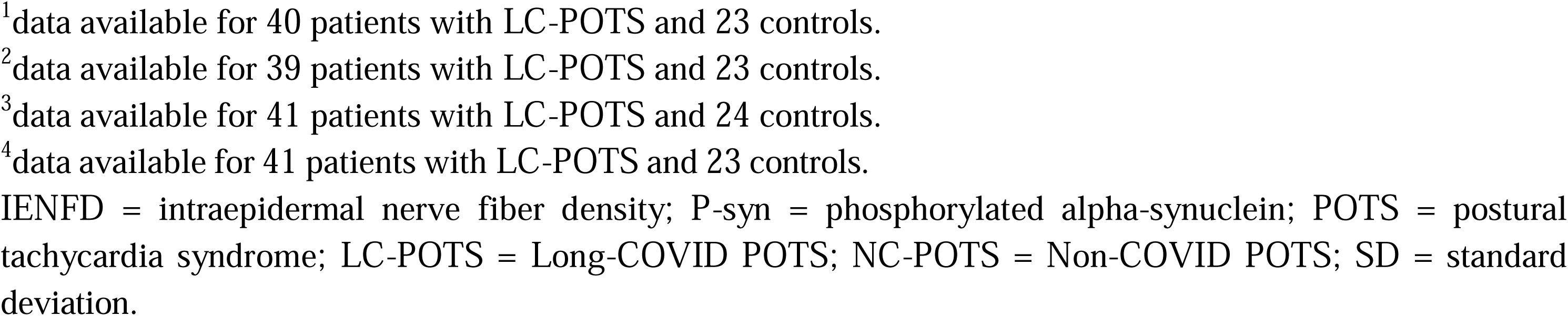
Skin Biopsy Results.

| Skin biopsies | LC-POTS (n = 41) | NC-POTS (n = 15) | Control (n = 24) | P-value | Post-hoc |
| --- | --- | --- | --- | --- | --- |
| **IENFD (fibers/mm): mean ± SD (% abnormal)** |  |  |  |  |  |
| Cervical<sup>1</sup> | 43.3 ± 6.0 (0) | 47.8 ± 8.1 (0) | 42.8 ± 5.1 (0) | 0.038 | NC-POTS vs. Control |
| Dist. Thigh<sup>1</sup> | 17.5 ± 4.4 (5) | 20.6 ± 4.0 (6.7) | 16.1 ± 3.3 (8.7) | 0.003 | NC-POTS vs. LC-POTS/Control |
| Dist. Leg<sup>2</sup> | 12.1 ± 3.4 (12.8) | 15.4 ± 3.8 (6.7) | 12.0 ± 4.1 (17.4) | 0.007 | NC-POTS vs. LC-POTS/Control |
| Any abnormal site, n (%)<sup>3</sup> | 5 (12.2) | 1 (6.7) | 4 (16.7) | 0.74 | — |
| Length dependent pattern, n (%)<sup>4</sup> | 5 (12.2) | 1 (6.7) | 4 (16.7) | 0.73 | — |
| **P-syn present: n (%)** |  |  |  |  |  |
| Cervical<sup>1</sup> | 2 (5) | 0 (0) | 0 (0) | 0.69 | — |
| Prox. Thigh<sup>2</sup> | 1 (2.5) | 0 (0) | 0 (0) | 1.0 | — |
| Dist. Leg<sup>2</sup> | 1 (2.5) | 0 (0) | 0 (0) | 1.0 | — |
| Abnormal any site, n (%)<sup>1</sup> | 2 (5) | 0 (0) | 0 (0) | 1.0 | — |

Comparisons across the three groups were performed using a one-way analysis of variance (ANOVA) for continuous variables. Comparisons for categorical variables were performed using Chi-square tests for $\geq 5$ or Fisher's exact tests for $n < 5$ . For variables with statistically significant overall group differences ( $p < 0.05$ ), post-hoc pairwise comparisons were conducted using Tukey's honestly significant difference (HSD) test following ANOVA. Statistical significance was defined as $p < 0.05$ .
<sup>1</sup>data available for 40 patients with LC-POTS and 23 controls.
<sup>2</sup>data available for 39 patients with LC-POTS and 23 controls.
<sup>3</sup>data available for 41 patients with LC-POTS and 24 controls.
<sup>4</sup>data available for 41 patients with LC-POTS and 23 controls.
IENFD = intraepidermal nerve fiber density; P-syn = phosphorylated alpha-synuclein; POTS = postural tachycardia syndrome; LC-POTS = Long-COVID POTS; NC-POTS = Non-COVID POTS; SD = standard deviation.

To explore the possibility that controls with abnormal IENFD (n=4) were responsible for the abnormalities seen on autonomic testing in the control group, we performed a secondary analysis after excluding these participants, resulting in broadly similar findings. The racial distribution differed slightly, with a higher percentage of White participants in both LC-POTS and NC-POTS compared with controls. HUT findings were also largely similar, although ΔRR became non-significant. TCD findings remained similar.

Serological markers were largely similar across the groups, with no significant differences for most measurements (Supplementary Table 6). There was a single control with a mildly abnormal HbA1C (5.7%). Exclusion of this participant did not alter the results.

## Discussion

The primary aim of our study was to perform deep phenotyping across cardiovascular, cerebrovascular, respiratory, and neurocutaneous domains to better understand the potential patterns of autonomic dysfunction in LC-POTS, and how these patterns may differ from those of NC-POTS. In our carefully selected cohort of previously healthy LC-POTS participants, we found evidence of autonomic dysfunction spanning sudomotor, cardiovagal, and sympathetic adrenergic domains, with high rates of hyperadrenergic responses on HUT, reduced IENFD, and low rates of cutaneous P-Syn deposition. However, we also found similar rates of some of these same abnormalities in NC-POTS and controls, despite controls having no pre-existing conditions or persistent symptoms after SARS-CoV-2 infection.

Fourty-three percent of LC-POTS and 44% of NC-POTS participants exhibited elevated upright NE levels, which, coupled with higher HR and SBP on HUT, is suggestive of sympathetic predominance. The subtype of “hyperadrenergic POTS” has been well-described in the literature,^9^ with patients often presenting with symptoms of panic and “adrenaline surge” sensations, tremulousness, hyperhidrosis, orthostatic hypertension, and tachycardia. In fact, this presentation has been estimated to occur in 30-60% of POTS patients^12^ and was the most common symptomatic presentation in our LC-POTS cohort. Mechanisms underlying a hyperadrenergic state after SARS-CoV-2 infection are not well understood. Studies have demonstrated increased resting muscle sympathetic nerve activity in healthy adults recovering from acute COVID-19,^13^ as well as increased low-frequency domain spectra of heart rate variability in those with LC,^14^ findings that suggest increased sympathetic activity in both the acute and chronic forms of COVID-19. In addition, LC has been associated with elevated cytokine levels, and animal studies have demonstrated increased sympathetic activation with direct infusion of inflammatory cytokines,^15^ suggesting a role of ongoing inflammation in the propagation of sympathetic hyperactivity. While other studies have demonstrated elevated cytokine levels in both LC^16^ and non-COVID POTS patients,^17^ we found no significant difference in cytokine levels across groups in our study, although only IL-6 and TNF-α were tested and only in a subset of participants.

11.6% of our LC-POTS participants had evidence of sympathetic adrenergic impairment, with higher rates (20%) in NC-POTS participants. Cardiovagal parasympathetic adrenergic dysfunction was observed in a smaller subset of our POTS cohort (9.3% in LC-POTS and 12.5% in NC-POTS). In a retrospective study of LC-POTS participants, 10% had abnormal HRVdb (n=10) on autonomic testing, rates similar to those observed in our cohort.^18^ Vagal impairment has been demonstrated in other chronic post-infectious illnesses, notably myalgic encephalitis/chronic fatigue syndrome (MC/CFS),^19^ and autopsy studies have demonstrated SARS-CoV-2 spike protein and nucleocapsid fragments in the dorsal motor nucleus of the vagus in LC,^20^ suggesting direct tissue invasion as one possible source of vagal impairment. In addition, vagal dysfunction may contribute to a pro-inflammatory state by means of inhibition of the vagal inflammatory reflex that mediates innate immune responses and systemic inflammation.^21^ Vagal nerve stimulation has been demonstrated to improve both the degree of postural tachycardia as well as symptom severity in patients with POTS without LC, supporting that vagal nerve dysfunction plays a role in the pathophysiology of POTS.^22^

Measures of cerebral blood flow on HUT were similar across groups, an unexpected finding. Other studies using unilateral right MCA doppler have demonstrated alterations in mean CBFv during HUT in both LC without POTS^23^ and non-COVID POTS.^24,25^ In contrast to control participants in these studies, controls in our study had all been previously infected with SARS-CoV-2, raising the possibility that some participants had post-viral changes that resulted in asymptomatic alterations in CBF regulation. Furthermore, our controls also demonstrated low but not insignificant rates of autonomic testing abnormalities, suggesting that some of them may not have been truly “healthy” despite a lack of documented medical comorbidities, lack of ongoing symptoms attributed to COVID, and absence of orthostatic symptoms during HUT. Having a second COVID-naïve control group could better answer this question. However, this is nearly impossible given the fact that most of the world population has been infected with SARS-CoV-2 at least once.

We found that participants in our LC-POTS cohort exhibited a significant degree of hypocapnia in both supine and upright positions during HUT. Possible mechanisms of hypocapnia in LC include hyperventilation (though mean supine and end-tilt RR in our cohorts were normal), undiagnosed pulmonary disease, and carotid body dysregulation. There is evidence that the carotid chemoreflex is amplified in LC, which may result in hyperventilation and reduced efficiency of breathing during exercise.^26^ Hypocapnia is a known cerebral vasoconstrictor, which in turn can result in decreased CBF.^27^ One study showed that both LC (POTS not specified) and non-COVID POTS exhibited a decrease in CBFv on HUT when compared to healthy, COVID-naïve, pre-pandemic controls.^23^ In this study, the CBFv decline in POTS participants was driven mainly by hypocapnia, while in LC, the CBFv decline was independent of CO_2._ It should be emphasized that this particular study used POTS and control groups recruited prior to the COVID pandemic, which may limit the generalizability of CBFv measurements in those with a history of SARS-CoV-2 infection, as in our study. Another study documented a decrease in CBFv during HUT in normocapnic POTS patients, suggesting that CO_2_ is not the only driver of altered CBFv in POTS.^24^ In our study, we did not find any significant correlations between CO_2_ and CBFv within either POTS groups or controls. Taken together, these findings suggest that although hypocapnia is common in LC-POTS, hypocapnia is not the only driver of CBFv changes during HUT. The high rates of hypocapnia seen in the control group were also unexpected, and could not be explained by tachycapnea. It is possible that asymptomatic recovery from post-viral changes contributed to these findings, however the exact mechanisms are unclear.

NIRS is a well-validated and non-invasive technique to measure peripheral and central tissue oxygen extraction. ^28^ NIRS uses the near-infrared region of the electromagnetic spectrum to allow for deeper penetration of light (up to 8 cm) into tissues to monitor changes in oxygen extraction^29,28^ Previous studies using NIRS have shown evidence of impaired cerebral^30^ and peripheral oxygen extraction^31^ in POTS. However, we found no significant differences in central tissue oxygen extraction between groups. Skeletal muscle rSO2 in the supine position was significantly lower in LC-POTS compared to controls, although the significance of this is unclear given that the skeletal muscle rSO2 during HUT did not differ between the groups. Overall, our results suggest that cerebral and peripheral oxygen extraction remained intact in our LC-POTS cohort.

Signs of SFN were common in our POTS cohort, with 12.2% of LC-POTS participants exhibiting reduced IENFD on skin biopsy and 39% exhibiting evidence of sudomotor dysfunction on Q-sweat. Our findings are consistent with a prior study in which 2/5 (40%) of LC-POTS patients had neuropathic findings on skin biopsy.^18^ Direct evidence of SFN has been described in both non-COVID POTS^32^ and LC without POTS.^23, 33^ In fact, reduced IENFD has been documented in 24-45% of patients with POTS prior to the COVID pandemic,^34^ although these findings have not always correlated with symptoms of SFN. In at least a subset of these patients, POTS is thought to result from partial sympathetic denervation leading to impaired vasoconstriction, reduced venous return, and compensatory tachycardia. Our results suggest that SFN, involving both the unmyelinated somatic and autonomic C fibers, is common among those with LC-POTS and could possibly contribute to the underlying physiology of POTS.

It should be noted that rates of IENFD abnormalities were lower in our NC-POTS group (6.7%), and relatively high in controls (16.7%), leading to no significant difference between groups. All LC-POTS participants with IENFD and/or Q-Sweat abnormalities reported symptoms of neuropathic pain after SARS-CoV-2 infection. In comparison, none of the controls reported neuropathic pain.

These results suggest that reduced mild reductions in IENFD may not correlate with symptoms of SFN in asympomatic infidividuals. It is possibile that the reduced IENFD in some controls could also have been due to post-viral damage from COVID, although all control participants had recovered fully from their acute COVID infection without symptoms suggestive of SFN. Excluding those controls with abnormal IENFD from the final analysis did not significantly alter our findings.

An unexpected finding of our study was that 5% of LC-POTS participants exhibited aggregates of P-syn on skin biopsy (one of these participants was previously reported in a prior case series^35^). We found no differences in autonomic test results or demographics between P-Syn positive and P-Syn negative LC-POTS participants. Both P-Syn positive LC-POTS participants were unvaccinated, which raises the possibility that COVID vaccination protected against P-Syn deposition, however it should be emphasized that these are only two cases and further study is warranted. The presence of P-Syn on skin biopsy has been reported in a small cohort of POTS patients in a single pre-COVID retrospective study.^36^ However, data on viral or other triggers were not available in this cohort.

Alpha-synuclein is a small protein expressed in monomeric, dimeric, and tetrameric formation in neurons of the central and peripheral nervous system, as well as red blood cells and other tissues, and is responsible for mediating the transport of synaptic vesicles.^37^ When phosphorylated, it undergoes a conformational change to a β-sheet structure that recruits additional monomers to form amyloid fibrils, resulting in the pathological aggregates of Lewy bodies, seen in Parkinson’s disease (PD) and Lewy body dementia (DLB) and glial cytoplasmic inclusions seen in multiple system atrophy (MSA). *In vitro* studies have demonstrated that the SARS-CoV-2 spike protein (S protein) interacts with α-syn to form strains with stronger seeding activity and neurotoxicity and that the aggregation and phosphorylation of α-syn were enhanced in cells overexpressing the S1 domain.^38^ A recent study reported the presence of a positive synuclein seed amplification assay (SAA) in olfactory mucosa samples of approximately one-third of individuals with persistent hyposmia after COVID,^39^ hypothesizing that SARS-CoV-2 infection may trigger local synuclein misfolding in the context of neuroinflammation. Notably, P-syn deposition was also seen in 3.3% of control participants, which is keeping with other studies showing similar rates of p-syn depotion in asymptomatic individuals.^10^

These results should be interpreted with an abundance of caution, and more robust pathobiological studies assessing the presence of P-Syn in the skin and other biospecimens, such as cerebrospinal fluid, in not only LC-POTS but other LC cohorts, are ultimately needed before these findings can be attributed to SARS-CoV-2. It is important to note that none of the participants in our cohort had skin biopsies prior to having COVID, so it is unclear whether COVID acted as the initial trigger of P-Syn deposition or whether P-Syn was present prior to COVID infection and increased the risk of developing autonomic complications from COVID-19. In addition, while cutaneous P-syn is assumed to be a very specific finding of synuclein-driven neurodegenerative disease, it is possible that SARS-CoV-2 may induce local neural inflammation, which may itself influence synuclein aggregation that is not directly associated with neurodegeneration. In support of this theory, we performed a follow-up biopsy set on one of our P-syn positive LC-POTS participants approximately one year after the baseline biopsy, finding that the participant was no longer P-syn positive, which also correlated with a significant improvement in POTS symptoms. Longitudinal tissue and biofluid studies are ultimately needed to help determine if P-syn deposition may be transient in other LC patients.

The strengths of our study include the carefully phenotyped, previously healthy cohort of LC-POTS participants with a confirmed SARS-CoV-2 trigger, many of whom were unvaccinated and infected early in the pandemic, an ideal cohort to study the effects of LC on autonomic dysfunction. This, combined with a well-phenotyped group of NC-POTS and the extensive battery of autonomic, neurophsysiologic, blood and tissue-based analyses provides a robust deep phenotyping comparison. The limitations of our study include the relatively small sample size of the NC-POTS and control groups, which may have limited statistical power to detect significant differences between groups.

This was due in part to the strict exclusion criteria combined with the burdensome testing regimen that limited recruitment. In addition, given the cross-sectional nature of this study, we were unable to determine whether either reduced IENFD or P-Syn seen on skin biopsy was present prior to SARS-CoV-2 infection. LC-POTS participants were also slightly older and had higher (BMI) compared to NC-POTS, which may have had some influence on autonomic testing results. Finally, all control participants had a history of COVID-19, so it is possible that some of the abnormalities found in the control group were the result of asymptomatic post-viral sequelae.

## Conclusions

Orthostatic tachycardia, sympathetic adrenergic hyperactivity, hypocapnia and other autonomic abnormalties were relatively common our in LC-POTS cohort, although may of these findings overlapped with NC-POTS and controls. These findings of have been reported separately in other POTS cohorts. However, ours is one of the few in which the etiology of POTS, SARS-CoV-2, was confirmed in all participants. While the presence of viral infection as a trigger of POTS has been supported for decades by various retrospective reports,^40^ the COVID-19 pandemic has offered a rare opportunity to validate this association with more rigorous methods and diagnostic testing. Prior studies have categorized POTS participants into various subtypes however our findings support the concept of multiple pathological processes in most patients with POTS triggered by SARS-CoV-2, a finding which can likely be extended to other viral pathogens. Indeed, the presence of many of the same abnormalities in our NC-POTS cohort confirmed similar mechanistic pathways in both POTS groups. In addition to sympathetic hyperactivity and small fiber neuropathy, other mechanisms implicated in POTS pathogenesis include hypovolemia, deconditioning, and autoimmunity.^41^ While most, if not all, of our participants reported reduced physical activity due to their ongoing symptoms, we did not quantify deconditioning or their volume status.

Finally, an asymptomatic decrease in cerebral perfusion and reduced IENFD were also seen in COVID-recovered controls, which may have been the result of sampling error due to small sample size, asymptomatic post-viral injury, or undiagnosed underlying medical conditions. Future studies should include a COVID naïve group control group when possible. Larger sample sizes are needed to confirm these findings and their relation to the pathophysiological mechanisms of autonomic dysfunction in COVID-19.

## Data Availability

All data produced in the present study are available upon reasonable request to the authors.

## Acknowledgements

The authors wish to thank Drs. Peter Novak, Linda van Campen, and Frans Visser for their assistance in TCD methodology.

## Contributions

Conception and design: NL, MM; analysis and interpretation of data: NL, MM, HBT, JS, RS; drafting of the manuscript or revising it critically for important intellectual content: NL, HBT, CG, PU, ML, SM, SJ, MM.

## Potential Conflics of Interest

Dr. Larsen has received grant funding from Dysautonomia International for the submitted work. Dr. Gibbons has stock options in CND Life Sciences, the company that performed the skin biopsy analysis for this study. Dr. Miglis has received grant funding from Dysautonomia International and the National Institutes of Health for the submitted work and is a member of the medical advisory board for Dysautonomia International (unpaid). Jannika Machnik, Jordan Seliger, Ruba Shaik, Maarten Lansberg, Srikanth Muppidi. PJ Utz, and Safwan Jaradeh have nothing to disclose.

