## Supplemental Tables for "Deep Phenotyping Long-COVID Postural Tachycardia Syndrome"

**Supplementary Table 1. Participant Demographics and Clinical Characteristics. stratified by COVID-19 infection timing (pre-omicron vs post-omicron), defined using a cutoff of December 1, 2021.**

| **Demographics** | **Pre-Omicron (n = 20)** | **Post-Omicron (n = 23)** | **P-value** |
| --- | --- | --- | --- |
| **Age, mean (SD)** | 35.5 (9) | 32.9 (8.9) | 0.34 |
| **Gender, n (%)** | | | |
| Male | 4 (20) | 1 (4.3) | 0.17 |
| Female | 16 (80) | 22 (95.7) |  |
| **Race, n (%)** | | | |
| White | 16 (80) | 18 (78.3) | 0.55 |
| White/Hispanic | 0 (0) | 0 (0) |  |
| Black | 1 (%) | 0 (0) |  |
| Asian | 1 (5) | 4 (17.4) |  |
| Native Hawaiian/Other Pacific Islander | 1 (5) | 0 (0) |  |
| Other | 0 (0) | 0 (0) |  |
| Not disclosed | 1 (5) | 1 (4.3) |  |
| **BMI (kg/m^2^), mean (SD)** | 27.5 (6.1) | 27.2 (7.4) | 0.55^a^ |
| **Time from infection to ANS testing (months), m**ean (SD) | 25.3 (15.7) | 15.5 (9.4) | 0.032^a^ |
| **COVID Severity, n (%)** | | | |
| Hospitalized | 15 (75) | 21 (91.3) | 0.22 |
| **Vaccinated at time of COVID infection, n (%)** | | | |
| Yes | 4 (20) | 19 (82.6) | 0.0001 |
| **UPSIT-40^1^** | | | |
| Mean (SD) | 34 (2.2) | 33.8 (3) | 1.0^a^ |
| Abnormal, n (%) | 5 (62.5) | 10 (83.3) | 0.34 |

Continuous variables are presented as mean (SD) and were compared between groups using independent t-tests for normally distributed variables or Mann–Whitney U^a^ tests for non-normally distributed variables, as determined by assessment of normality. Categorical variables are presented as counts (percentages) and were compared using Chi-square tests for expected cell counts ≥ 5 or Fisher’s exact tests for expected counts < 5.

^1^data available only for 8 pre-omicron and 12 post-omicron participants.

*Abbreviations:* BMI = body mass index; ANS = autonomic nervous system; SD = standard deviation; COVID-19 = coronavirus disease 2019; Pre-Omicron = infection prior to December 1, 2021; Post-Omicron = infection on or after December 1, 2021.

**Supplementary Table 2. Autonomic Testing results stratified by COVID-19 infection timing (pre-omicron vs post-omicron), defined using a cutoff of December 1, 2021.**

| **Variables** | **Pre-Omicron (n = 20)** | **Post-Omicron (n = 23)** | **P-value** |
| --- | --- | --- | --- |
| **HRVdb** | | | |
| Mean (SD) | 23.4 (7.3) | 22.2 (9.6) | 0.63 |
| Abnormal, n (%) | 1 (5) | 2 (8.7) | 1.0 |
| **Valsalva Ratio** | | | |
| Mean ± SD | 2.1 (0.4) | 2 (0.4) | 0.52 |
| Abnormal, n (%) | 1 (5) | 1 (4.3) | 1.0 |
| Parasympathetic dysfunction, n (%)^*^ | 1 (5) | 3 (15) | 0.61 |
| **Phase IIL abnormal, n (%)** | 1 (5) | 3 (13.04) | 0.61 |
| **Phase IV abnormal, n (%)** | 1 (5) | 2 (8.7) | 1.0 |
| **Sympathetic adrenergic dysfunction, n (%^**^** | 2 (10) | 3 (13.0) | 1.0 |
| **Q-SWEAT: mean (SD)** | | | |
| Forearm^1^ | 0.7 (0.4) | 0.7 (0.6) | 0.76 |
| Forearm abnormal, n (%)^1^ | 0 (0) | 2 (9.1) | 0.49 |
| Prox. Thigh^2^ | 0.8 (0.5) | 0.7 (0.5) | 0.57 |
| Prox. Thigh abnormal, n (%)^2^ | 2 (10.5) | 3 (13.6) | 1.0 |
| Dist. Thigh^2^ | 0.6 (0.4) | 0.4 (0.3) | 0.15 |
| Dist. Thigh abnormal, n (%)^2^ | 3 (15.8) | 3 (13.6) | 1.0 |
| Foot^2^ | 0.4 (0.3) | 0.4 (0.5) | 0.82 |
| Foot: abnormal, n (%)^2^ | 4 (21.0) | 4 (18.2) | 1.0 |
| Abnormal any site, n (%)^2^ | 6 (33.3) | 9 (40.9) | 0.87 |
| Length dependent, n (%)^2^ | 4 (21.0) | 4 (18.1) | 0.78 |
| **Norepinephrine (pg/mL): mean (SD)**^4^ | | | |
| NE Supine | 297.9 (139) | 363.1 (292.1) | 0.36 |
| NE Upright | 574.6 (206.7) | 605.4 (387.4) | 0.76 |
| NE Delta | 276.6 (199.6) | 282.6 (243.1) | 0.93 |
| Hyperadrenergic, n (%)^***^ | 9 (47.4) | 8 (61.5) | 0.78 |
| **CASS: mean (SD)**^5^ | | | |
| Sudomotor: mean (SD) | 0.7 (1.3) | 0.5 (0.9) | 0.86^a^ |
| Cardiovagal: mean (SD) | 0.2 (0.7) | 0 (0) | 0.29^a^ |
| Sympathetic: mean (SD) | 0.1 (0.3) | 0.2 (0.6) | 0.42 |
| Total: mean (SD) | 0.4 (0.9) | 0.4 (0.8) | 0.95 |

Continuous variables are presented as mean (SD) and were compared between groups using independent t-tests for normally distributed variables or Mann–Whitney U^a^ tests for non-normally distributed variables, as determined by assessment of normality. Categorical variables are presented as counts (percentages) and were compared using Chi-square tests or Fisher’s exact tests, as appropriate. Statistical significance was defined as a two-sided p < 0.05.**^*^**Cardiovascular parasympathetic dysfunction defined by abnormal Valsalva ratio and/or abnormal HRDB. ^**^Sympathetic Adrenergic Dysfunction defined by abnormal phase IIL and/or abnormal phase IV. ^***^Hyperadrenergic response defined by upright NE>600 and/or >3-fold increase in NE from supine to upright.

^1^data available only for 18 pre-omicron and 22 post-omicron participants.

^2^data available only for 19 pre-omicron and 22 post-omicron participants.

^3^data available only for 19 pre-omicrons and 21 post omicron participants.

^4^data available only for 7 pre-omicron and 11 post-omicron participants.

^5^data available only for 7 pre-omicron and 11 post-omicron participants.

*Abbreviations:* HRVdb = heart rate variability during deep breathing; SD = standard deviation; NE = norepinephrine; Q-sweat = quantitative sudomotor axon reflex testing.

**Supplementary Table 3. Head Up Tilt Results, stratified by COVID-19 infection timing (pre-omicron vs post-omicron), defined using a cutoff of December 1, 2021.**

|  | **Pre-Omicron (n = 20)** | **Post-Omicron (n = 23)** | **P-value** |
| --- | --- | --- | --- |
| **HR (bpm)** | | | |
| Supine mean (SD) | 74.2 (11.7) | 75.8 (11.9) | 0.65 |
| Max Tilt mean (SD) | 114.7 (19.9) | 119 (23.9) | 0.52 |
| End Tilt mean (SD) | 101.8 (18.5) | 110 (28.5) | 0.25 |
| Δ(Max-tilt - Supine) | 40.5 (15.4) | 43.2 (21.3) | 0.92^a^ |
| Δ(End-tilt - Supine) | 27.6 (13) | 34.3 (27.5) | 0.31 |
| **Systolic BP (mmHg)** | | | |
| Supine | 125.4 (17.2) | 125.8 (13) | 0.94 |
| Max Tilt | 149.1 (22.4) | 139.7 (22.6) | 0.18 |
| End Tilt | 133.6 (16.6) | 123.3 (21.1) | 0.081 |
| Δ(Max-tilt - Supine) | 23.6 (19.5) | 13.9 (22.8) | 0.14 |
| Δ(End-tilt - Supine) | 8.2 (15.8) | -2.5 (20.4) | 0.062 |
| **ETCO_2_ (mmHg)^1^** | | | |
| Supine | 29.6 (5.6) | 26.5 (7.2) | 0.19^a^ |
| Min Tilt | 23.1 (6.4) | 17.9 (6.8) | 0.022^a^ |
| End Tilt | 25.4 (6.8) | 19 (6.9) | 0.19^a^ |
| Δ(Supine - Min tilt) | 6.5 (3.4) | 8.7 (6.4) | 0.20 |
| Δ(Supine - End-tilt) | 4.3 (4.4) | 7.5 (7.2) | 0.092 |
| **RR (breaths/min)^1^** | | | |
| Supine | 15.3 (5.7) | 14.8 (4.2) | 0.77 |
| Max Tilt | 18.5 (5.2) | 20.9 (6.1) | 0.38^a^ |
| End Tilt | 16.7 (5.6) | 17.3 (5.5) | 0.89^a^ |
| Δ(Max-tilt - Supine) | 3.2 (3.4) | 6 (5.9) | 0.074 |
| Δ(End-tilt - Supine) | 1.4 (4.1) | 2.5 (6) | 0.52 |
| **Arterial Oxygen Saturation (SaO_2_ %)^2^** | | | |
| Supine | 93.9 (18.4) | 98.1 (1.9) | 0.63 |
| Min Tilt | 96.4 (2.9) | 92 (17.1) | 0.81 |
| End Tilt | -2.4 (17.6) | 6.1 (16.1) | 0.17 |
| Δ(Supine - Min tilt) | 96.9 (3) | 97.5 (2.2) | 0.75 |
| Δ(Supine - End-tilt) | -2.9 (18) | 0.6 (1.4) | 0.35 |
| **NIRS Right Forehead (rSO_2_ %)**^3^ | | | |
| Supine | 78.7 (2.5) | 76.9 (6) | 0.80^a^ |
| Min Tilt | 74.7 (3.4) | 72.1 (5.9) | 0.10 |
| End Tilt | 76.6 (2.6) | 73.4 (6.1) | 0.036 |
| Δ(Supine - Min tilt) | 4 (2.9) | 4.8 (2.5) | 0.082^a^ |
| Δ(Supine - End-tilt) | 2.1 (1.9) | 3.5 (3) | 0.070 |
| **NIRS Left Forehead (rSO_2_ %)3** | | | |
| Supine | 77.6 (2.9) | 76.2 (5.9) | 0.79^a^ |
| Min Tilt | 69.9 (13.8) | 71.7 (5.4) | 0.75^a^ |
| End Tilt | 74.8 (3.6) | 72.4 (5.3) | 0.10 |
| Δ(Supine - Min tilt) | 7.7 (13.5) | 4.5 (2.4) | 0.99^a^ |
| Δ(Supine - End-tilt) | 2.7 (2.3) | 3.8 (2.7) | 0.40^a^ |
| **NIRS Calf (rSO_2_)**^2^ | | | |
| Supine | 73.2 (5.4) | 72.2 (4.6) | 0.57 |
| Min Tilt | 64.6 (9.7) | 67 (8.8) | 0.54 |
| End Tilt | 65.0 (9.8) | 67.9 (9.4) | 0.45 |
| Δ(Supine - Min tilt) | 8.5 (9.3) | 5.2 (8.7) | 0.22 |
| Δ(Supine - End-tilt) | 8.2 (9.4) | 4.4 (9.2) | 0.15 |

Values are presented as mean (SD). Continuous variables are presented as mean (SD) and were compared between groups using independent t-tests for normally distributed variables or Mann–Whitney U^a^ tests for non-normally distributed variables, as determined by assessment of normality. Statistical significance was defined as a two-sided p < 0.05.

^1^data available only for 19 pre-omicron and 20 post-omicron participants.

^2^data available only for 19 pre-omicron and 21 post-omicron participants.

^3^data available only for 18 pre-omicron and 21 post-omicron participants.

*Abbreviations:* HR = heart rate; SBP =systolic blood pressure; ETCO₂, end-tidal carbon dioxide; NIRS, near-infrared spectroscopy; RR = respiratory rate; rSO₂, regional oxygen saturation; SD = standard deviation.

**Supplementary Table 4. Table 4. Head Up Tilt Transcranial Doppler Results**, **stratified by COVID-19 infection timing (pre-omicron vs post-omicron), defined using a cutoff of December 1, 2021.**

| **TCD Variable (cm/s)** | **Pre-Omicron (n = 16)** | **Post-Omicron (n = 16)** | **P-value** |
| --- | --- | --- | --- |
| **Baseline Left MCA mean (SD)** | | | |
| Mean vCBF (cm/s) | 60.2 (12.4) | 62.3 (13.2) | 0.92^a^ |
| Peak SV (cm/s) | 89.2 (17.1) | 94.7 (20.9) | 0.42 |
| Peak DV (cm/s)^1^ | 46.1 (5.3) | 39.5 (8.6) | 0.091^a^ |
| PI | 0.8 (0.1) | 0.8 (0.1) | 0.49 |
| **End Tilt Left MCA** mean (SD) | | | |
| Mean vCBF (cm/s)^2^ | 46 (13.7) | 47.9 (15.9) | 0.73 |
| Peak SV (cm/s)^2^ | 71 (21.3) | 78.8 (24.7) | 0.35 |
| Peak DV (cm/s)^1^ | 35.6 (7.8) | 31.4 (10.2) | 0.32 |
| PI^2^ | 0.9 (0.2) | 1 (0.4) | 0.59^a^ |
| **Delta left MCA mean (SD)** | | | |
| Mean vCBF (cm/s)^2^ | 14.4 (10.6) | 14.5 (14) | 0.99 |
| Peak SV (cm/s)^2^ | 18.2 (13.1) | 15.9 (14.4) | 0.64 |
| Peak DV (cm/s)^1^ | 10.5 (10.3)^1^ | 8.1 (13.1) | 0.66 |
| PI^2^ | -0.1 (0.3) | -0.2 (0.4) | 0.59^a^ |
| **Baseline Right MCA mean (SD)** | | | |
| Mean vCBF (cm/s)^3^ | 58 (12.6) | 62.5 (12.5) | 0.33 |
| Peak SV (cm/s)^3^ | 87.9 (15) | 95.4 (21.1) | 0.27 |
| Peak DV (cm/s)^5^ | 45.4 (5.8) | 41.6 (9.7) | 0.31 |
| PI^3^ | 0.9 (0.2) | 0.8 (0.2) | 0.39^a^ |
| **End Tilt Right MCA mean (SD)** | | | |
| Mean vCBF (cm/s)^4^ | 48.3 (10.4) | 51.8 (13.9) | 0.45 |
| Peak SV (cm/s)^4^ | 75.5 (16.7) | 85.6 (22.2) | 0.18 |
| Peak DV (cm/s)^5^ | 37.2 (6.6) | 34.6 (10) | 0.52 |
| PI^4^ | 0.8 (0.1) | 1 (0.4) | 0.45^a^ |
| **Delta Right MCA mean (SD)** | | | |
| Mean vCBF (cm/s)^4^ | 10.6 (7.5) | 10.7 (11.8) | 0.98 |
| Peak SV (cm/s)^4^ | 13.8 (7.4) | 9.8 (15.5) | 0.38 |
| Peak DV (cm/s)^5^ | 8.3 (5.7) | 7 (10.2) | 0.74 |
| PI^4^ | 0 (0.2) | -0.2 (0.4) | 0.12^a^ |

Continuous variables are presented as mean (SD) and were compared between groups using independent t-tests for normally distributed variables or Mann–Whitney U^a^ tests for non-normally distributed variables, as determined by assessment of normality. Statistical significance was defined as a two-sided p < 0.05.*

^1^data available for 11 post-omicron and 8 pre-omicron participants.

^2^data available for 16 post-omicron and 15 pre-omicron participants.

^3^data available for 15 post-omicron and 15 pre-omicron participants.

^4^data available for 15 post-omicron and 13 pre-omicron participants.

^5^data available for 11 post-omicron and 7 pre-omicron participants.

*Abbreviations:* TCD = transcranial Doppler; MCA = middle cerebral artery; FV = flow velocity; PSV = peak systolic velocity; PDV =peak diastolic velocity; PI = pulsatility index; SD = standard deviation.

TCD = transcranial Doppler; MCA = middle cerebral artery; SV = systolic velocity; DV = diastolic velocity; PI = pulsatility index; SD = standard deviation; vCBF = mean cerebral blood flow velocity.

**Supplementary Table 5. Table 5. Skin Biopsy Results, stratified by COVID-19 infection timing (pre-omicron vs post-omicron), defined using a cutoff of December 1, 2021.**

| **Skin biopsies** | **Pre-Omicron (n = 19)** | | **Post-Omicron (n = 22)** | **P-value** |
| --- | --- | --- | --- | --- |
| **IENFD (fibers/mm): mean ± SD (% abnormal)** | | | | |
| Cervical^1^ | 43.29 (7.07) | 43.44 (4.7) | | 0.32 |
| Dist. Thigh^1^ | 18.37 (5.48) | 16.48 (2.39) | | 0.15 |
| Dist. Leg^2^ | 12.12 (3.97) | 12.13 (2.78) | | 0.83 |
| Abnormal any site, n (%)^3^ | 3 (13.6%) | 2 (10.5%) | | 1.0 |
| Length dependent pattern, n (%)^3^ | 3 (13.6%) | 2 (10.5%) | | 1.0 |
| **P-syn present: n (%)** | | | | |
| Cervical^1^ | 1 (4.5%) | 1 (5.6%) | | 1 |
| Prox. Thigh^4^ | 1 (4.5%) | 0 (0%) | | 1 |
| Dist. Leg^4^ | 0 (0%) | 1 (5.9%) | | 0.44 |
| Abnormal any site, n (%)^1^ | 1 (4.5%) | 1 (5.6%) | | 1 |

Continuous variables are presented as mean (SD) and were compared between groups using independent t-tests for normally distributed variables. Categorical variables are presented as counts (percentages) and were compared using Chi-square tests or Fisher’s exact tests, as appropriate. Statistical significance was defined as a two-sided p < 0.05.*

¹data available for 18 pre-omicron and 22 post-omicron participants.

²data available for 18 pre-omicron and 21 post-omicron participants.

³data available for 19 pre-omicron and 22 post-omicron participants.

⁴data available for 17 pre-omicron and 22 post-omicron participants.

*Abbreviations:* IENFD =intraepidermal nerve fiber density; P-syn = phosphorylated alpha-synuclein.

**Supplementary Table 6. Serological and laboratory markers across participants with LC POTS, NC POTS, and controls.**

| **Variable** | **LC POTS (n = 19)** | **NC POTS (n = 17)** | **Controls (n = 19)** | **P-value** |
| --- | --- | --- | --- | --- |
| **Complete blood count** | | | | |
| WBC | 7 (2.2) | 6.9 (2) | 5.8 (1.7) | 0.12 |
| Hemoglobin | 13.4 (1) | 13.2 (1.3) | 13.4 (1.5) | 0.99^a^ |
| Hematocrit | 40.6 (3.2) | 40.1 (3.7) | 40.9 (3.8) | 0.96^a^ |
| Platelet count | 290.9 (56.1) | 276.5 (69.1) | 251.3 (47.7) | 0.099^a^ |
| RBC | 4.5 (0.5) | 6.3 (6.7) | 4.6 (0.5) | 0.81^a^ |
| MCV | 90.6 (4.7) | 88.7 (5.4) | 89.4 (5) | 0.56 |
| MCH | 30 (1.8) | 29.2 (2.4) | 29.4 (1.9) | 0.39^a^ |
| MCHC | 33.1 (0.7) | 31.2 (5.9) | 32.8 (1.2) | 0.24^a^ |
| RDW | 12.7 (0.7) | 12.2 (2.4) | 12.8 (0.9) | 0.83^a^ |
| **Differential counts** | | | | |
| Neutrophils (%) | 60.2 (10.5) | 60.1 (11) | 60.6 (8.7) | 0.99 |
| Lymphocytes (%) | 28.3 (8) | 29.3 (9) | 29.2 (7.3) | 0.93 |
| Monocytes (%) | 7.2 (1.9) | 7.2 (2) | 6.7 (1.6) | 0.64 |
| Eosinophils (%) | 3.4 (5.5) | 2.4 (1.7) | 2.7 (2.6) | 0.82^a^ |
| Basophils (%) | 0.6 (0.3) | 0.8 (0.4) | 0.6 (0.3) | 0.18^a^ |
| Immature granulocytes (%) | 0.3 (0.1) | 0.3 (0.1) | 0.3 (0.1) | 0.88^a^ |
| NRBC (%) | 0 (0) | 0 (0) | 0 (0) | — |
| Neutrophils (absolute) | 4.4 (2.1) | 4.2 (1.7) | 3.6 (1.4) | 0.55^a^ |
| Lymphocytes (absolute) | 1.9 (0.5) | 1.9 (0.4) | 1.7 (0.5) | 0.31 |
| Monocytes (absolute) | 0.5 (0.2) | 0.5 (0.2) | 0.4 (0.1) | 0.062 |
| Eosinophils (absolute) | 0.2 (0.4) | 0.2 (0.1) | 0.2 (0.2) | 0.95^a^ |
| Basophils (absolute) | 0 (0) | 0.1 (0) | 0 (0) | 0.14^a^ |
| Immature granulocytes (absolute) | 0 (0) | 0 (0) | 0 (0) | 0.25^a^ |
| NRBC (absolute) | 0 (0) | 0 (0) | 0 (0) | — |
| **Immune profiling / lymphocyte subsets** | | | | |
| CD3+ pan T cells (%) | 73.1 (7.9) | 74.6 (5.8) | 75.1 (7.7) | 0.37^a^ |
| CD3+CD4+ (%) | 44.7 (5.5) | 44.5 (8.8) | 43.2 (8.6) | 0.82 |
| CD3+CD8+ (%) | 25.8 (6.3) | 26.5 (5.5) | 28.5 (6.2) | 0.38 |
| CD4/CD8 ratio | 1.8 (0.5) | 1.8 (0.7) | 1.6 (0.5) | 0.37 |
| CD3+ T cells (absolute) | 1438.1 (401.9) | 1293.8 (326.7) | 1253.2 (373.2) | 0.29 |
| CD4+ T cells (absolute) | 869.4 (203.7) | 770.4 (255.8) | 722.5 (259.7) | 0.18 |
| **Coagulation markers** | | | | |
| D-dimer | 0.5 (0.1) | 0.3 (NA) | 0.4 (0.1) | 0.26 |
| **Electrolytes/acid-base** | | | | |
| Sodium | 138.8 (2.5) | 138.9 (1) | 138.5 (1.7) | 0.63^a^ |
| Potassium | 4 (0.3) | 4.1 (0.2) | 4.1 (0.4) | 0.91^a^ |
| Chloride | 104.7 (3) | 103.1 (1.4) | 98.1 (21.3) | 0.066^a^ |
| CO_2_ | 24 (2.2) | 24.7 (2.8) | 25.4 (2) | 0.22 |
| Anion gap | 10.3 (2.4) | 11.1 (2.8) | 10.1 (1.6) | 0.45 |
| Calcium | 9.5 (0.3) | 9.7 (0.5) | 9.3 (0.3) | 0.043 |
| **Renal function** | | | | |
| Urea nitrogen | 12.4 (3.3) | 10.8 (2.5) | 11.6 (2.9) | 0.33^a^ |
| Creatinine | 0.7 (0.1) | 0.7 (0.1) | 0.8 (0.2) | 0.19 |
| eGFR | 112.2 (11.6) | 113.8 (19.5) | 113.5 (15.4) | 0.69^a^ |
| **Liver function/proteins** | | | | |
| Total bilirubin | 0.6 (0.5) | 0.4 (0.2) | 0.6 (0.2) | 0.10^a^ |
| AST | 21.4 (7.1) | 24.8 (8.8) | 27.4 (9.7) | 0.33^a^ |
| ALT | 23.1 (12.2) | 22.2 (16.1) | 24.1 (20.5) | 0.78^a^ |
| Alkaline phosphatase | 64.5 (18.4) | 61.4 (17.5) | 60.4 (16) | 0.77 |
| Albumin | 8.9 (18.3) | 4.6 (0.3) | 4.6 (0.2) | 0.88^a^ |
| Total protein | 7.2 (0.8) | 7.4 (0.4) | 7.4 (0.4) | 0.81^a^ |
| Globulin | 3.1 (1.2) | 2.8 (0.3) | 2.8 (0.3) | 0.84^a^ |
| **Metabolic markers** | | | | |
| Glucose | 101.5 (31.5) | 89.9 (11.1) | 94.5 (12.5) | 0.41^a^ |
| Hemoglobin A1c | 5.4 (1) | 5.1 (0.8) | 5.2 (0.3) | 0.22^a^ |
| **Cardiovascular/nutritional markers** | | | | |
| NT-proBNP | 54.1 (17.5) | 57 (16.2) | 59 (15.8) | 0.84 |

Continuous variables are presented as mean (SD). Group comparisons were performed using one way ANOVA or Kruskal Wallis^a^ testing as appropriate. The total number of participants included for each measurement varied because values below the lower limit of detection and measurements that were not performed were excluded from analyses

*Abbreviations:* ALT = alanine aminotransferase; AST = aspartate aminotransferase; CO₂ = carbon dioxide; eGFR = estimated glomerular filtration rate; LC-POTS = long COVID postural tachycardia syndrome; MCH = mean corpuscular hemoglobin; MCHC = mean corpuscular hemoglobin concentration; MCV = mean corpuscular volume; NC-POTS = non-COVID postural tachycardia syndrome; NRBC = nucleated red blood cells; NT-proBNP = N-terminal pro B-type natriuretic peptide; RBC = red blood cells; RDW = red cell distribution width; WBC = white blood cells.
